# Enrichment of Rare and Uncommon Neanderthal Polymorphisms in Autistic Probands and Siblings

**DOI:** 10.1101/2023.10.27.23297672

**Authors:** Rini Pauly, Layla Johnson, F. Alex Feltus, Emily L. Casanova

**Affiliations:** Biomedical Data Science and Informatics Program, Clemson University, Clemson, SC 29634, USA; Department of Psychology, Loyola University, New Orleans, New Orleans, LA, 70118, USA, USA; Department of Genetics and Biochemistry, Clemson University, Clemson, SC 29634, USA; Center for Human Genetics, Clemson University, Clemson, SC 29634, USA

## Abstract

*Homo sapiens* and Neanderthals underwent hybridization during the Middle/Upper Paleolithic age, culminating in retention of small amounts of Neanderthal-derived DNA in the modern human genome. In the current study, we address the potential roles genic Neanderthal single nucleotide polymorphisms (SNP) may be playing in autism susceptibility using data from the Simons Foundation Powering Autism Research (SPARK) and Genotype-Tissue Expression (GTEx) databases. We have discovered that rare and uncommon variants are significantly enriched in both European- and African-American autistic probands and their unaffected siblings compared to race-matched controls. In addition, we have identified 51 SNPs (p51) significantly enriched in European-American cases of autism, 13 of which fall within autism-associated genes, as well as 1 SNP in African-American probands. In addition, SNPs within the p51 network display significant linkage disequilibrium with one another, indicating they may more often be co-inherited in autism. These results strongly suggest Neanderthal-derived DNA is playing a significant role in autism susceptibility across major populations in the United States.

## BACKGROUND

DNA evidence taken from human remains from the Middle Pleistocene indicate that anatomically modern humans (AMH) and other archaic humans underwent multiple introgression events [*1*]. Of those archaic humans, *Homo neanderthalensis* (Neanderthals) have received the most attention, providing the most fossil material, and occupying the position as our closest known cousin on the hominin tree of life. It has been estimated that Eurasian-derived populations have approximately 2% Neanderthal DNA, which was acquired during introgression events occurring shortly after AMH migrated out of Africa [*2,3*]. These hybridization events occurred somewhere between 47-65 thousand years ago (kya) [*4*]. A subset of Europeans later immigrated back into Africa approximately 20 kya, bringing some of this Neanderthal ancestry with them, such that modern Africans have a small but measurable amount of Neanderthal DNA from the event [*5*].

With the recent sequencing of multiple archaic human genomes, there has been growing interest concerning the influence of archaic human-derived alleles on modern health [*6,7*]. With regards to Neanderthal-derived variants, previous groups have identified positive selection on genes relating to immune function, skin and hair pigmentation, physiological responses to high altitude conditions, aspects of metabolism, hypercoagulation, and propensity for depression [*6,8*].

In general, dosage-sensitive genes are tightly conserved and resistant to such introgression events [*9,10*]. Most genes involved in brain development follow this dosage-sensitive pattern and have tended to be resistant to introgression [*11*]. In support of this, Srinivasan et al. (2016) found a depletion of Neanderthal-derived variants within autism- and other brain-related genes in the general population. On the other hand, other studies have reported a number of non-synonymous and other single nucleotide variants within neural genes that have been implicated in the condition [*3,12*].

Additional research involving non-clinical populations has identified strong links between certain brain and skull morphologies and enrichment of Neanderthal DNA [*13,14*]. Specifically, enrichment is associated with reduced globularity in the skull shape of modern populations, a finding mildly reminiscent of the elongated skull morphology characteristic of Neanderthal and other archaic crania [*14*]. Enrichment of Neanderthal DNA is also associated with enhanced neural connectivity within visual processing systems, particularly between the intraparietal sulcus (IPS) and the occipital cortex and fusiform gyrus, and decreased connectivity within the default mode (social) network [*13,15*].

Importantly, many of these same connectivity patterns are recapitulated in autism, which is a major impetus for the current work. For instance, underconnectivity within the default mode network is consistently reported in autism [*16*]. Meanwhile, increased connectivity within visual processing networks is also a commonly reported feature. Keehn et al. [*17*] tested autistic children^1^ using a visual search task in which this group tends to excel. They identified an “island of sparing” in autism reflected in increased connectivity within occipital regions and enhanced connectivity of these visual processing areas to the frontal lobes. Autistic people also often have cognitive strengths in areas such as mathematics. Iuculano et al. [*19*] found that during numerical problem solving, autistic children had greater activation in the fusiform gyrus and occipital lobes compared to neurotypical children, again suggesting that visual processing modalities are an area of strength in autism. Interestingly, many of these features are shared by non-clinical groups with high Neanderthal DNA content.

In light of this evidence, in the current study we addressed whether Neanderthal DNA is enriched in autistic people and their siblings compared to non-autistic controls. We accessed whole exome sequencing (WES) for autistic probands and unaffected siblings from the Simons Foundation Powering Autism Research (SPARK) Database [*20*] for comparison against individuals in the Genotype-Tissue Expression (GTEx) database [*21*]. Significant enrichment in the autistic probands and their sibs was driven by rare and uncommon Neanderthal-derived variants, which suggests weak but ongoing purifying selection towards removal of these single nucleotide polymorphisms (SNP) from the human genome.

## MATERIALS AND METHODS

An assemblage of Neanderthal-derived SNPs was provided by the Sankararaman laboratory and has been previously published [*7,22*]. Only European-specific SNPs were used in the current study as no participants were of self-reported Asian background. The authors applied for and received access to WES from the SPARK dataset (Clemson University IRB2018-235), which is a collection of extensive genotype and clinical data on autistic individuals and their unaffected siblings [*20*]. Samples have been collected from a wide range of collection sites within the US and can be found on the SFARI website (see: https://www.sfari.org/resource/spark/). Only individuals of self-reported “white” (European-American) and African-American backgrounds were used in these analyses; any participant listed as “admixed” was removed. Autistic individuals with potential confounding genetic and environmental factors were also excluded from the study. These conditions include: birth or pregnancy complications, premature birth, fetal alcohol syndrome, and cognitive delays due to exposure or medical condition (see Supplementary Table 1 for basic demographics). European- and African-American controls were accessed from the GTEx Project (Clemson University IRB2022-0589) for comparison.

**Table 1.**
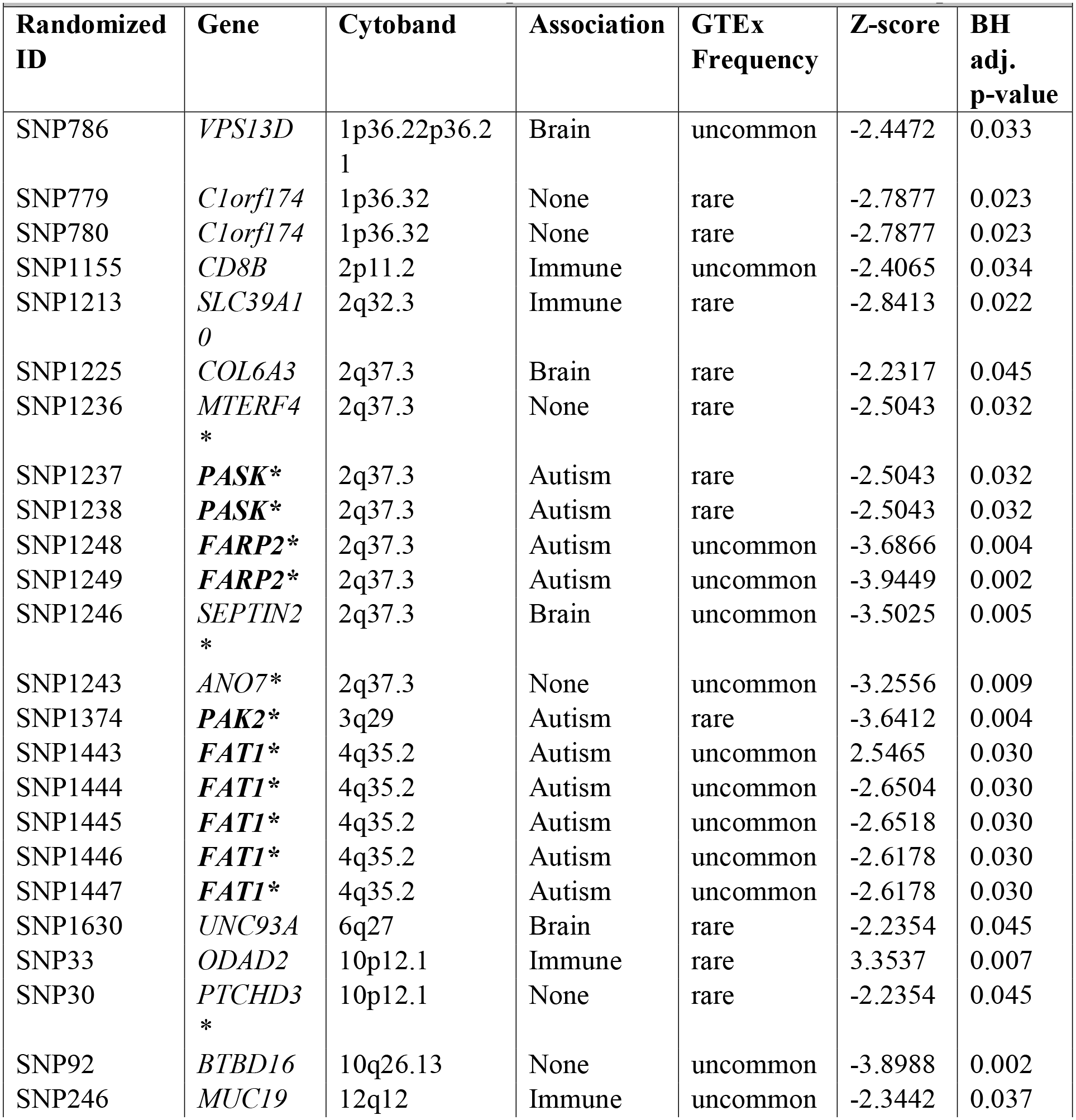
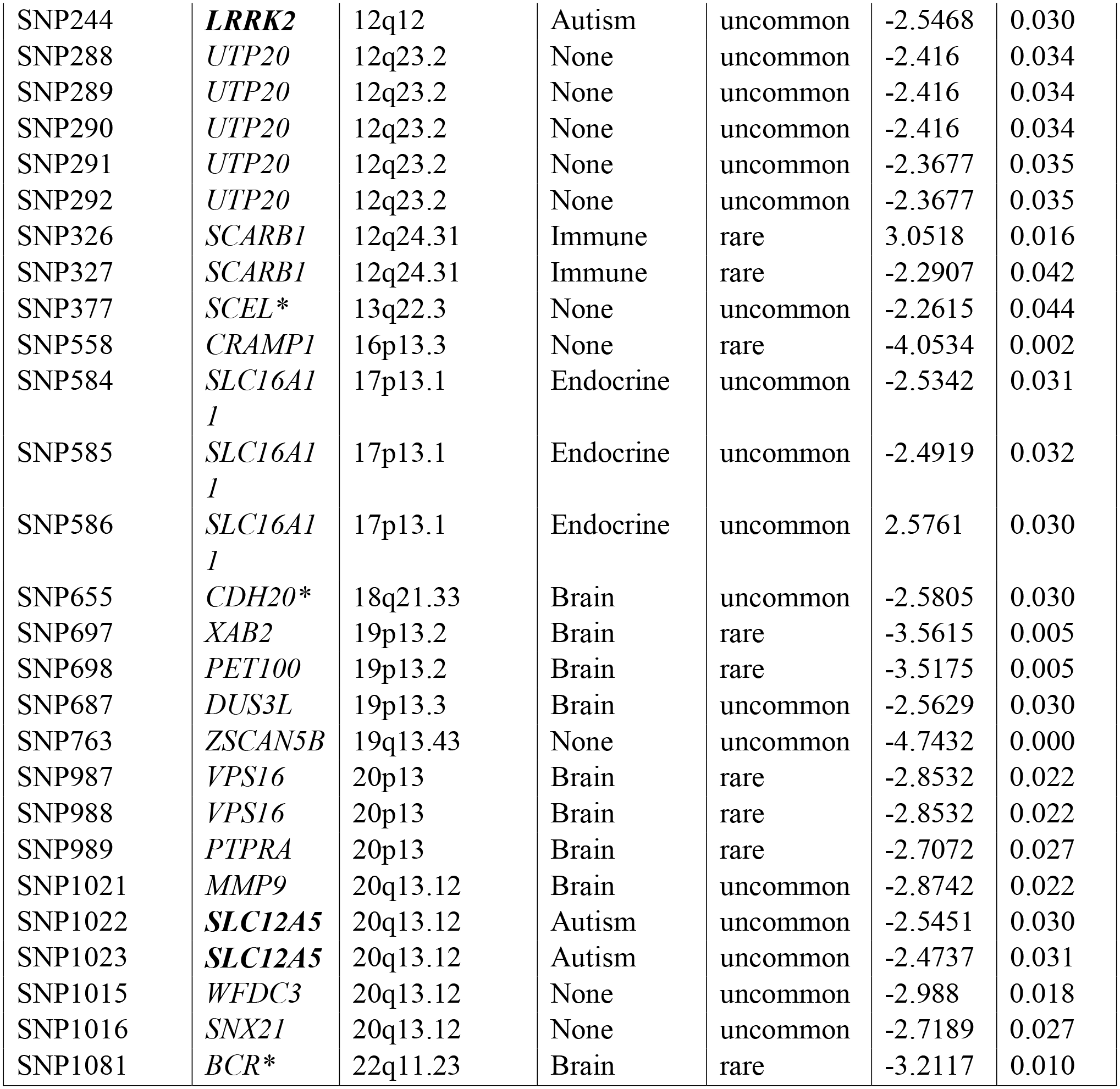
P51 SNPs Enriched in European-American SPARK Affected Sample. List of 51 SNPs (p51) in 35 genes significantly enriched in European-American SPARK affected participants compared to GTEx controls. Thirteen SNPs within 6 different genes with known association to autism are marked in bold. A further 12 genes are expressed in brain, 5 involved in immune system regulation, and 1 with the endocrine system. Seventeen SNPs are also enriched, though not significantly, in the African-American SPARK group (marked with an asterisk), 10 of which are targeting autism-associated genes.

Both European- and African-American allele frequencies were determined using the GTEx database. While numbers of European-American exomes were sufficient (GTEx N = 715, SPARK affected = 2,316, SPARK unaffected siblings = 360), the number of African-American SPARK affected and GTEx individuals were comparatively small (SPARK N = 75, GTEx N = 103), which limited our analysis of this latter subset. All SNPs with missing data (which occurred solely within the SPARK datasets) were removed from the analyses. This left a total of 1,288 SNPs for comparison across all five groups.

To process the genotype calls from SPARK and GTEx VCFs, the individual genotypes were converted into a binary call format (bcf) using bcftools (see: https://github.com/samtools/bcftools). Subsequently, the bcf files were transformed into genotype matrices using the JVARKIT tools developed by Pierre Lindenbaum. The JVARKIT tools can be found at the following GitHub repository (see: https://github.com/lindenb/jvarkit/).

European-American SNPs were categorized as “rare” (*<* 1%), “uncommon” (1% ≥ *x* ≤ 5%), and “common” (*>* 5%), according to their frequency within the GTEx dataset. Meanwhile, within the African-American groups, SNPs were broken down into “rare” (*<* 1%) and “non-rare” (1% ≥) according to their GTEx frequency. Across all groups, homozygous ancestral (non-Neanderthal) SNPs were assigned a value of “0,” heterozygous Neanderthal SNPs were given a value of “1,” and homozygous Neanderthal alleles assigned the value of “2.” Raw NeanderScores were then calculated for each individual according to the average of these values and were subsequently broken down into the rare, uncommon, common, and non-rare categories for their respective groups.

A list of 51 European-American SNPs (p51) was derived by selecting all SNPs whose frequencies were 1% or higher in the SPARK affected group compared to race-matched controls. From there, we performed two-proportion Z-tests with BH multitest correction, producing the p51 SNP list that appears to have significant association with the autism group. In order to then produce the weighted Cytoscape networks, we calculated which of the p51 SNPs were in linkage disequilibrium (LD) with one another, using an *r*^*2*^ cutoff of 0.01 due to the rarity of the SNPs studied. This resulted in a list of 50 SNPs with measurable LD to build the network in Cytoscape (see Supplementary Table 2 for matched list of sources and targets). These networks were then analyzed using the Network Analyzer function in Cytoscape in order to derive the degrees (first degree neighbors) for each SNP by group. In addition, degree averages were statistically analyzed by group.

### Statistics

NeanderScores across total SNPs and common SNPs followed a normal distribution and therefore parametric statistics were used (i.e., ANOVA), with standard *post hoc* comparison. Meanwhile, due to skewed distributions of rare and uncommon SNPs in the European-Americans and rare and non-rare SNPs in African-Americans, nonparametric analyses such as Mann-Whitney U and Kruskal-Wallis were used, with Games-Howell *post hoc* comparison when appropriate. A student’s *t* test was also used to compare the number of degrees per node within the p51 linkage LD Cytoscape network analysis. For paired comparison of nonparametric data, a Wilcoxon signed-rank test was used and for binomial data (e.g., presence/absence), chi-square and odds ratios. For covariates such as Group by Sex, ANCOVAs were used. Analyses were conducted and plots produced using R and Cytoscape, the former with the *ggplot2* package.

## RESULTS

A raw “NeanderScore” is an estimate of the Neanderthal DNA content within a given person’s genome, calculated based on the average number of Neanderthal-derived SNPs [*7,22*]. In the current study, all NeanderScores were calculated using genic content. Total NeanderScores differed significantly across groups, with NeanderScores considerably greater in the European-Americans compared to African-Americans as expected [*F*(4, 3564) = 889.317, *p <* 0.0001, *η*^2^ = 0.500] (Fig. 1A) (*5*). However, according to *posthoc* analysis, total NeanderScores did not differ between experimental and control groups within the European- and African-American subsets respectively.

**Fig. 1.**
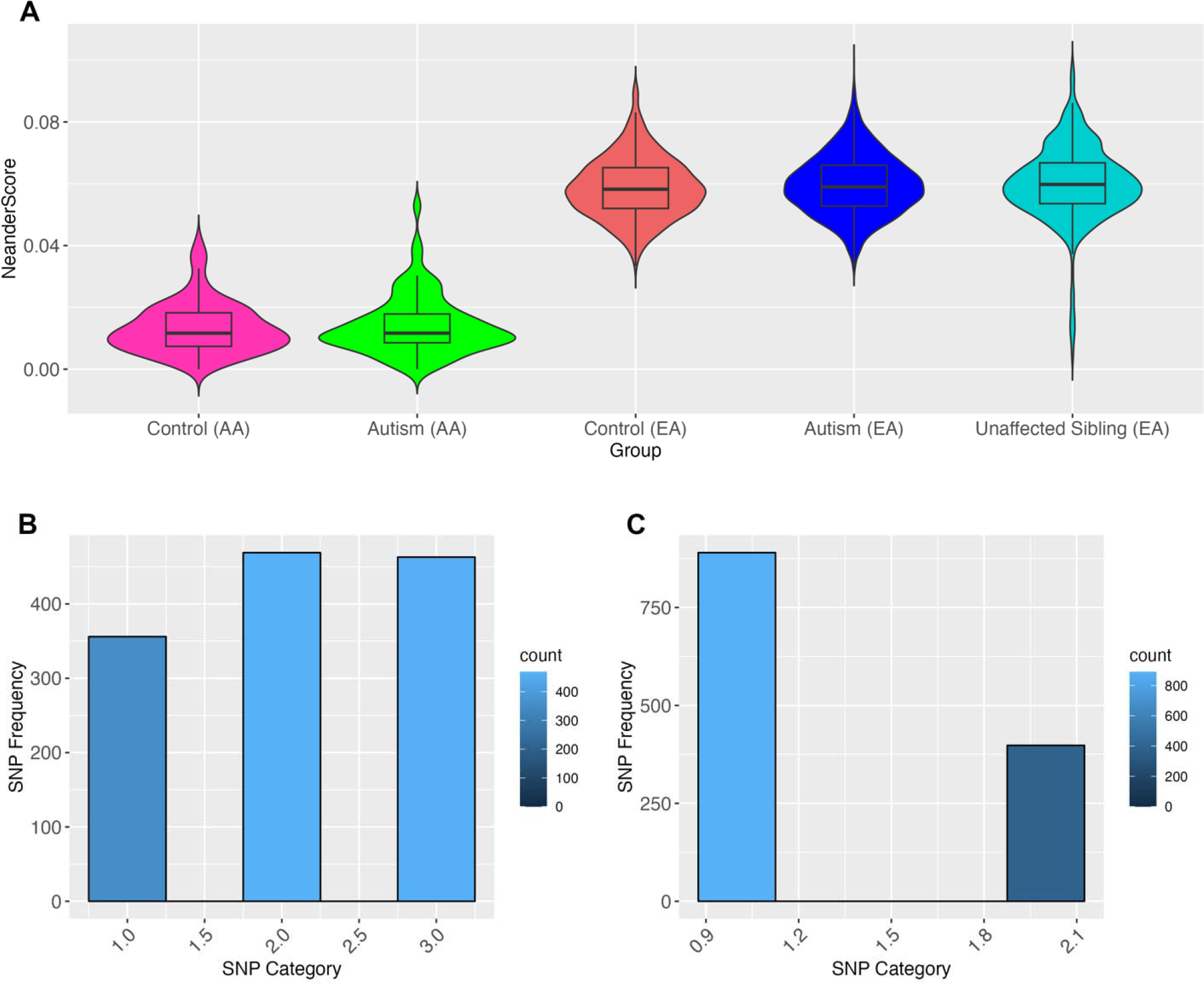
(A) Total NeanderScores across all groups. (EA = European-American, AA = African-American.) (B) Frequency of SNPs in European-American controls, divided into rare, uncommon, and common categories. (C) Frequency of SNPs in African-American controls, divided into rare and non-rare categories.

Rare SNPs occur, by definition, in less than 1% of the population, while uncommon SNPs occur between 1-5% and common in 5% or more [*23*]. Within the European-American groups, SNPs were broken down into these three categories (rare, uncommon, and common) according to their frequency within the GTEx European-American group and analyzed separately (Fig. 1B). Meanwhile, within the African-American groups, due to smaller participant numbers and lower Neanderthal content, SNPs were broken down into “rare” and “non-rare” categories (Fig. 2C). According to these approaches, rare and uncommon SNPs within the European-American GTEx group composed approximately two thirds of genic SNPs (rare = 29%, uncommon = 34%), suggesting weak purifying selection against Neanderthal DNA within European-American genic regions, a finding that matches previous research [*24*] (Fig. 1B). Meanwhile, the majority of genic SNPs (69%) within the African-American genome would be considered rare as a result of low overall Neanderthal content in African-derived populations (Fig. 1C).

**Fig. 2.**
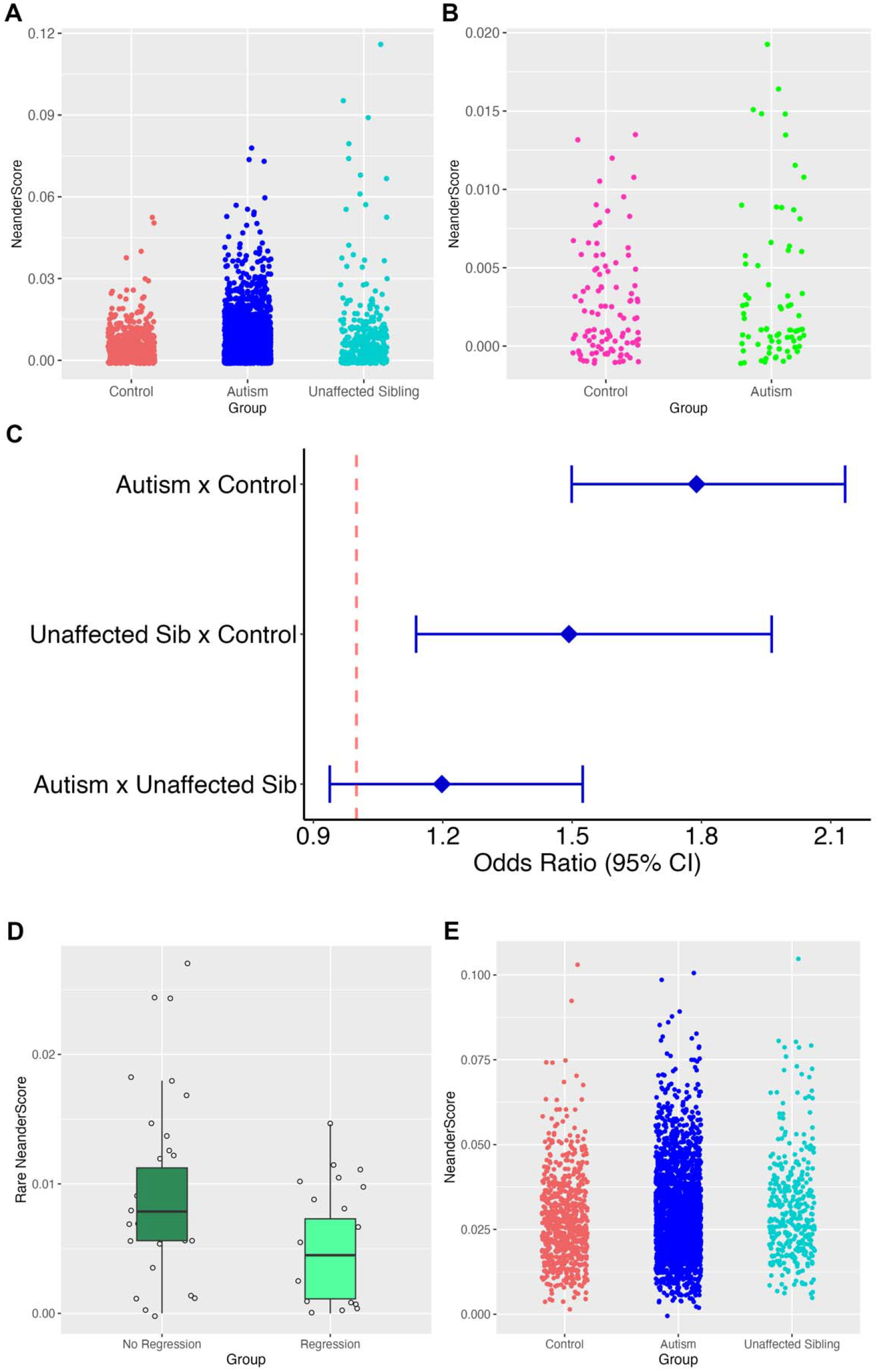
(A) Rare NeanderScores across European-American groups. (B) Rare NeanderScores across African-American groups. (C) Forest plot displaying odds ratios of rare SNPs across European-American experimental groups compared to controls. (D) Rare NeanderScores in African-American autistic probands grouped by the presence vs. absence of language regression. (E) Uncommon NeanderScores across European-American groups.

Although total NeanderScores did not differ between GTEx and SPARK groups, there was a dramatic enrichment of rare SNPs in all SPARK groups compared to matched controls (European-American: *H* = 61.012, *df* = 2, *p* = 5.641e-14; African-American: *W* = 2021.500, *p* = 4.880e-8) (Fig. 2A-B). According to Games-Howell *post hoc* comparison, European-American SPARK affected and unaffected siblings significantly differed from controls (*p* = 2.590e-12 and 9.809e-7, respectively), but they did not significantly differ from one other (*p* = 0.135). Approximately 56% of European-American controls had one or more rare genic SNPs, while 70% of SPARK affected individuals had rare Neanderthal variants [*OR* = 1.79, *95% CI* (1.50, 2.13), *z* = 6.45, *p* = 6.867e-11] (Fig. 2C). A similar enrichment was seen in the European-American SPARK unaffected siblings compared to matched GTEx controls [*OR* = 1.49, *95% CI* (1.14, 1.96), *z* = 2.90, *p* = 0.003], while the SPARK groups did not differ from one another [*OR* = 1.2, *95% CI* (0.94, 1.52), *z* = 1.45, *p* = 0.140]. European-American SPARK and GTEx groups also significantly differed from one another in rare homozygous Neanderthal content (*H* = 22.52, *df* = 2, *p* = 1.288e-5), with both experimental groups housing significantly more rare homozygous Neanderthal SNPs than controls following Games-Howell *posthoc* comparison (p = 2.085e-5-4.350e-2). Lastly, although sex by group were significant predictors of rare SNP enrichment [*F* (2, 3385) = 2.637, *p* = 0.026], group comparisons did not survive multitest correction, suggesting sex was not playing a measurable role in rare SNP content (*p* = 0.645-0.908).

We then performed a similar analysis on the African-American groups, finding– much like the pattern seen in the European-American analysis– the number of individuals with GTEx-defined rare SNPs was significantly greater in the affected group compared to controls (93.3% vs. 82.5% respectively; *OR* = 2.95, *95% CI* [0.99, 10.68], *z* = 1.94, *p* = 0.042) (Fig. 2B). In addition, there was no significant effect of group by sex in African-Americans [*F* (1, 174) = 0.463, *p* = 0.497].

In order to address whether rare SNP enrichment in the SPARK groups was a reflection of biased mode of selection, we flipped the analysis and defined rarity according to SNP frequency within the European-American affected group. We found that SNPs that were rare in the European-American SPARK group were similarly rare in matched controls, indicating there is a subset of GTEx-defined rare SNPs enriched in SPARK that appears to be related to phenotype susceptibility– however, not all rare SNPs are implicated (*W* = 836050.000, *p* = 0.674).

Following these analyses, we investigated additional clinical associations connected with rare NeanderScores. Within the African-American SPARK group, scores were significantly associated with language regression. Specifically, individuals without language regression had rare NeanderScores double those with regression (*W* = 379, *p* = 0.002) (Fig. 2D). Meanwhile, there were no direct clinical associations between rare NeanderScores in the European-American SPARK affected group. However, there was a significant interaction between seizures and sex as a predictor of scores [*F* (0.0004, 0.173) = 5.839, *p* = 0.016]. In particular, SPARK females with epilepsy had rare NeanderScores almost double that of other subgroups. It should be noted rare NeanderScores did not differ between autistic individuals with and without intellectual disability (ID), suggesting Neanderthal DNA is playing a role in susceptibility in autistic people with idiopathic ID and those without (*W* = 353865.5, *p* = 0.903). The same trend held true for multiplex vs. simplex cases (*t* = 0.452, *df* = 2213, *p* = 0.651).

Next, we analyzed uncommon and common SNPs. Much like rare SNPs, uncommon SNPs (1-5%) also differed across European-American groups [*F* (2, 3388) = 10.569), *p* = 2.7e5] (Fig. 2E). Both SPARK affected and unaffected groups exhibited significant enrichment compared to controls in Games-Howell *post hoc* comparison (affected: *p* = 7.8e-4; unaffected: *p* = 6e-5). We then used an ANOVA to assess enrichment of common SNPs across European-Americans, which differed from one another [*F* (2, 3388) = 3.136, *p* = 0.044]; however, in standard *posthoc* comparison only SPARK unaffected significantly differed from GTEx (*p* = 0.036) and this is driven by unaffected female siblings [ANCOVA (Group x Sex): *F* (2, 3385) = 3.30, p = 0.037]. In contrast, while the African-American SPARK affected group was enriched for rare SNPs, the opposite pattern held true for non-rare SNPs, the latter enriched in controls (*W* = 4606, *p* = 0.0285).

We next investigated specific SNP enrichment in SPARK affected groups compared to controls. We selected all SNPs whose frequencies were 1% or higher in SPARK affected cases as a starting list and performed two-proportion Z-tests. Following Benjamini-Hochberg (BH) multitest correction, this left 51 rare and uncommon SNPs (labeled “p51” here for the sake of brevity) in 35 genes significantly enriched within the European-American SPARK affected group (Table 1) and only a single SNP in the gene, *RAD1*, in the African-American SPARK group where small participant numbers reduced detection power (*Z* = -3.3917, *BH adj. p* = 0.021). Interestingly, although they did not reach significance, 17 of these SNPs were at least 1% higher in frequency in the African-American SPARK group compared to controls– with some as high as 3-4 percentage points– suggesting with larger group numbers these SNPs may rise to significance. Ten of the 17 SNPs in the African-American SPARK group are targeting autism-associated genes (Table 1) and using a Wilcoxon signed-rank test for paired comparison, this pattern significantly differed between the African-American groups, suggesting the association with autism genes is not coincidence though these SNPs did not survive multitest correction (*W* = 77, *z* = 2.201, *p* = 0.014).

We extended this analysis by investigating LD in the p51 SNPs. Using these results, we built interaction networks with LD pairs in Cytoscape and found that the average number of first degree neighbors (degrees) was significantly greater in the autism group compared to controls (18.560 vs. 14.520; *t* = 2.050, *df* = 98, *p* = 0.043), indicating increased LD within this network in autism (Fig. 3A-B). A number of these SNPs are forming notable interaction hubs and have unusually high degrees of connectivity in the autism group compared to controls (see Fig. 3C). These results suggest not only are these individual SNPs enriched in autism, they are forming an extended network and may be inherited together in this clinical group.

**Fig. 3.**
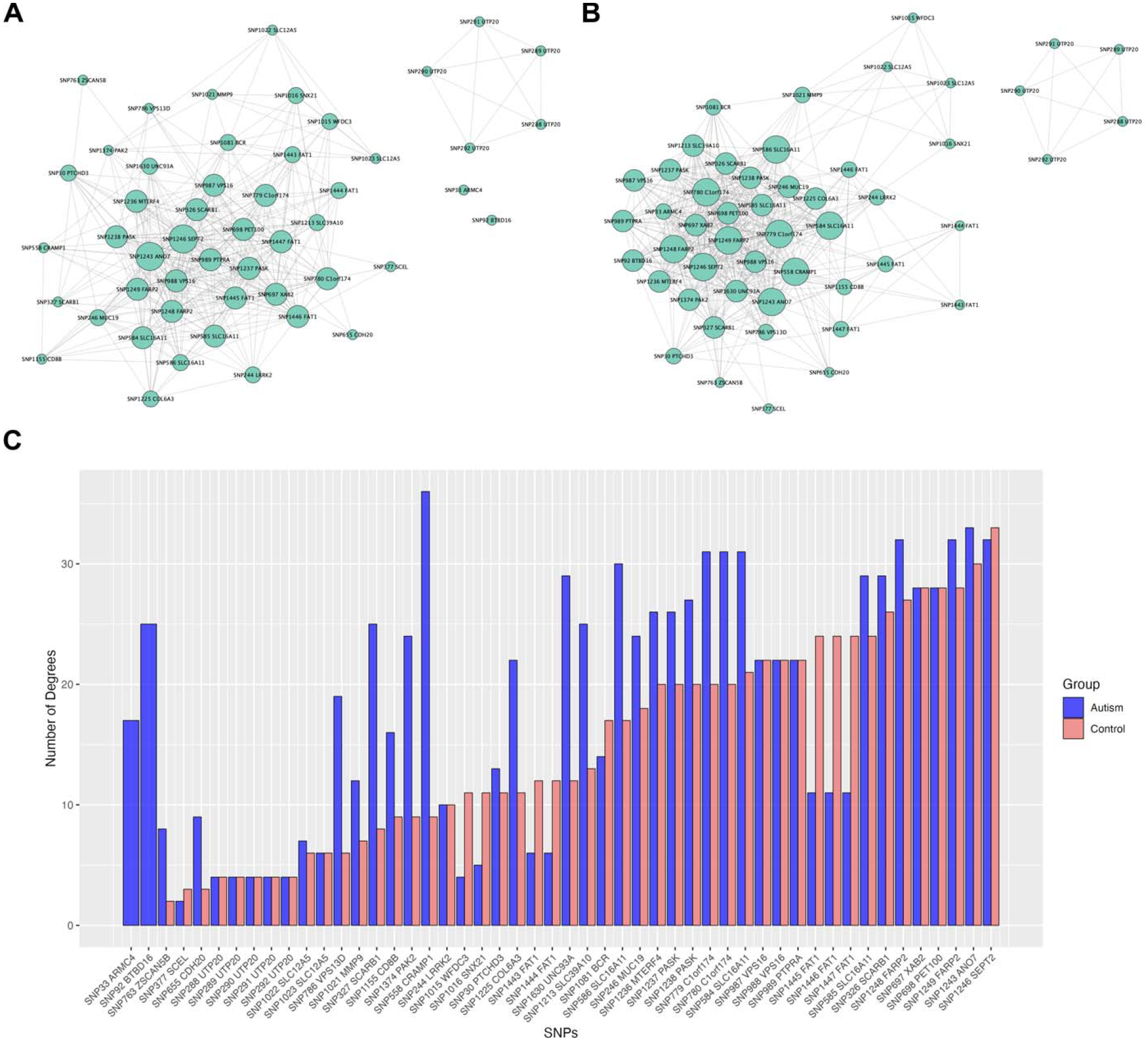
(A) P51 linkage disequilibrium in the control network showing reduced average connectivity (14.520 degrees) compared to the autism network (18.560 degrees) in (B). Nodes are weighted in size by number of degrees (direct neighbors). (C) Bar graph displaying degrees of connectivity per SNP by group. Note: All SNPs are listed with their randomized ID number in addition to the genes in which they reside.

Finally, we addressed phenotype correlations with enrichment of p51 in the European-American SPARK affected group. There was a significant interaction between multiplex vs. simplex status and the presence or absence of language regression [*F* (1, 2210) = 7.366, *p* = 0.007]. Specifically, individuals with language regression from multiplex families had significantly higher p51 NeanderScores than multiplex cases without language regression (5.644% vs. 3.596%, *p* = 0.046). Meanwhile, simplex language regression vs. no regression subgroups differed little from each other (3.873% vs. 4.240%, *p* = 0.826). However, all autism subgroups significantly differed from controls (*p <* 0.001), indicating these SNPs are susceptibility factors across European-American cases in general.

## DISCUSSION

Neanderthals experienced a prolonged genetic bottleneck, leading to greater retention of nonsynonymous mutations within their dwindling population, suggesting some of these variants may still be represented in the human genome today [*25*]. Juric et al. [*26*] found that these weakly deleterious variants, though retained in Neanderthals as a result of small population size, have been under purifying selection once they entered the *H. sapiens* background with access to a larger population. In support, Wei et al. [*24*] reported Neanderthal variants are significantly depleted in the modern genome relative to alleles matched for frequency and linkage disequilibrium.

Here we report an enrichment of rare and uncommon Neanderthal-derived polymorphisms in European- and African-American autistic people and their unaffected siblings. The low frequency of these SNPs, along with their clinical association, suggests they are weakly deleterious and under purifying selection. This is not, however, accompanied by an enrichment in overall Neanderthal content in SPARK groups, suggesting these low-frequency SNPs are playing measurable, complex roles in autism susceptibility.

In addition to studying large groups of SNPs, we analyzed individual polymorphisms, finding a total of 51 SNPs spread across 35 genes that are significantly enriched, co-occurring more frequently together in European-American SPARK affected cases (Table 1, Fig. 3). Six of these genes, represented by 13 SNPs, have known associations with autism. For instance, both deletions and duplications of 4q35.2 involving the *FAT1* gene have been found in autism, while animal models show the gene is expressed both within embryonic nervous system and postnatal cerebellum within the plasma membrane, soma, neurites, and postsynaptic densities [*27–29*]. Meanwhile, deletions of 2q37, including *FARP2* and *PASK* genes, have been found in autism, leading to their downregulated expression [*30,31*]. *FARP2*, in particular, has been shown to regulate dendrite morphogenesis [*32*]. Meanwhile, 3q29 microdeletion syndrome, which includes the gene, *PAK2*, is associated with autism and its haploinsufficieny leads to synaptic cytoskeletal impairments [*33,34*]. The neuronal chloride transporter, *SLC12A5* (KCC2), is important for the excitatory-to-inhibitory GABA perinatal shift and has been implicated in autism, epilepsy, and schizophrenia [*35, 36*]. Finally, *LRRK2* is the major candidate gene within copy number variants at the 12q12 locus associated with autism [*37*]. While these 6 genes have been implicated in autism, a further 12 genes in this SNP list are expressed in brain tissue, 5 in immune system regulation, and 1 in endocrine system regulation, suggesting future avenues of research [*38,39*].

Only a single significant SNP was identified in African-American probands in this study– specifically within the gene, *RAD1*. Although this gene is primarily implicated in DNA repair, it also interacts with topoisomerase 1 (Top1), which has previously been implicated in autism with its ability to regulate transcription of long nervous system-related genes [*41, 42*].

Although most studies on autism genomics focus on the deleterious nature of variants, there is the possibility some of these autism-associated Neanderthal SNPs have been under weak positive selection. In support, recent studies have identified genetic variants implicated in both autism and high intelligence [*40*]. Meanwhile, autistic people often perform better on tests of fluid intelligence than neurotypicals [*43*]. The variable penetrance of these Neanderthal variants for autism, as evidenced by high NeanderScores in unaffected siblings, suggests a means for their retention. In further support, studies on the cognitive ability of unaffected siblings show sibs tend to have higher performance IQ scores relative to verbal IQ – a pattern very similar to affected siblings and different from neurotypicals [*44*]. In addition, families of students studying disciplines like math, physics, and engineering are more likely to have autistic family members than students studying the humanities, suggesting an extended cognitive phenotype exists [*45*].

Of potential relevance to this topic, paleoarcheologists have identified an Upper Paleolithic Revolution beginning around 50 kya, which was a remarkable renaissance in the ways AMH made tools, traveled and engaged in trade, used and produced artistic materials, and structured their living habitats, such as dividing the home into food preparation, discard, cooking, and sleeping areas [*46*]. Interestingly, this time period roughly coincides with hybridization between *H. sapiens* and Neanderthals, suggesting hybridization may have been a stimulus for cognitive change, one which may continue to influence intellectual ability and susceptibility to neurodevelopmental conditions in modern humans. This has been hypothesized before, although from a sociocultural rather than a genetics perspective (*47*). Further work is needed to address these possibilities.

### Limitations

Despite strong clinical associations reported here, analyzing large numbers of SNPs gives the viewer only a gross perspective of potential roles Neanderthal-derived SNPs may be playing in autism susceptibility. While a subset of variants may be playing combinatory roles in susceptibility, there is much noise surrounding this genic signal and the current work would benefit in future from greater magnification of networks of SNPs driving these results.

In addition, there is the possibility the SPARK patient population is a biased collection of samples and subpopulations with unique Neanderthal genotypes may be over-represented compared to GTEx. However, if this were the case, it is difficult to explain: 1) Why inheritance of a subset of rare and uncommon SNPs in autism and their families would be preferenced in this scenario across both European- and African-American populations, 2) Why clinical and demographic characteristics, such as language regression, seizures, multiplex vs. simplex status, and sex, would associate with patterns in rare NeanderScores in clinical groups, and 3) Why there is enrichment overlap across the two clinical groups concerning the p51 alleles. Further research is needed to explore these possibilities but are beyond the current scope of this work.

## Conclusion

This is the first study to provide strong evidence for the active role of a subset of rare and uncommon Neanderthal-derived alleles in autism susceptibility in both European- and African-American populations. We hope this research will lead to further investigation into the ongoing influences of ancient hybridization between *H. sapiens* and Neanderthals in brain development, human intelligence, and overall human health, as well as spur work into additional clinical resources for this complex population.

## Data Availability

All deidentified data is available upon reasonable request to the authors. Full datasets can be accessed following approval via the Simons Foundation and the GTEx Project, respectively.

## Acknowledgments

We wish to thank the Sankararaman laboratory for providing the assemblage of Neanderthal-derived SNPs of the modern human genome. We also wish to thank the Simons Foundation and the Genotype-Tissue Expression Project for use of their respective datasets in the current study.

## Funding

N/A

## Author Contributions

RP curated the data and performed preliminary analyses. LJ produced the graphics for the study. FAF helped conceive the overall design of the study, provided data access, and mentored work at the Clemson site. ELC initially conceived of the study, performed the majority of data analysis, and mentored work at the Loyola site.

## Competing Interests

The authors declare no competing interests.

## Supplementary Materials

Due to data confidentiality required for genetic samples, full data may be acquired through the SPARK and GTEx Databases. Information on basic demographics and the Cytoscape network analysis can be found in the “Supplementary_Materials.docx” file on MP’s website.

## Footnotes

1 Within the current manuscript, the authors have elected to use identity-first language in accordance with current research on preferences of autism stakeholders (*18*).

## Notes

### Competing Interest Statement

The authors have declared no competing interest.

### Funding Statement

This study did not receive any funding.

### Author Declarations

Use of the SPARK database was approved by the Clemson University IRB (IRB2018-235). Use of the GTEx Project database was approved by the Clemson University IRB (IRB2022-0589).

