## Supplementary Materials for "Enrichment of Rare and Uncommon Neanderthal Polymorphisms in Autistic Probands and Siblings"

**Supplementary Tables**

**Supplementary Table 1.** Basic demographics of experimental and control groups in the study.

| **Group** | **N** | **Males** | **Females** |
| --- | --- | --- | --- |
| Controls (EA) | 715 | 472 | 243 |
| Autism (EA) | 2,316 | 1,822 | 494 |
| Unaffected Sibs (EA) | 360 | 180 | 180 |
| Controls (AA) | 103 | 71 | 32 |
| Autism (AA) | 75 | 63 | 12 |

**Supplementary Table 2.** Source, Target, and *r^2^* data by group for Cytoscape analysis. SNP pairs with extremely high *r^2^* scores (e.g., <0.7) occur within the same cytobands and are likely forming a haplotype.

| **Group** | **Source** | **Target** | ***r^2^*** |
| --- | --- | --- | --- |
| Autism | SNP1015 WFDC3 | SNP1016 SNX21 | 0.977301954 |
| Autism | SNP1015 WFDC3 | SNP1021 MMP9 | 0.175451852 |
| Autism | SNP1016 SNX21 | SNP1021 MMP9 | 0.17369825 |
| Autism | SNP1015 WFDC3 | SNP1022 SLC12A5 | 0.146204803 |
| Autism | SNP1016 SNX21 | SNP1022 SLC12A5 | 0.148997738 |
| Autism | SNP1021 MMP9 | SNP1022 SLC12A5 | 0.898957312 |
| Autism | SNP1015 WFDC3 | SNP1023 SLC12A5 | 0.139078383 |
| Autism | SNP1016 SNX21 | SNP1023 SLC12A5 | 0.141685328 |
| Autism | SNP1021 MMP9 | SNP1023 SLC12A5 | 0.737998378 |
| Autism | SNP1022 SLC12A5 | SNP1023 SLC12A5 | 0.818193884 |
| Autism | SNP1021 MMP9 | SNP1081 BCR | 0.011909234 |
| Autism | SNP1155 CD8B | SNP1225 COL6A3 | 0.015706053 |
| Autism | SNP1213 SLC39A10 | SNP1225 COL6A3 | 0.01304026 |
| Autism | SNP1081 BCR | SNP1236 MTERF4 | 0.0109552 |
| Autism | SNP1213 SLC39A10 | SNP1236 MTERF4 | 0.020926384 |
| Autism | SNP1225 COL6A3 | SNP1236 MTERF4 | 0.062353815 |
| Autism | SNP1081 BCR | SNP1237 PASK | 0.0109552 |
| Autism | SNP1213 SLC39A10 | SNP1237 PASK | 0.020926384 |
| Autism | SNP1225 COL6A3 | SNP1237 PASK | 0.062353815 |
| Autism | SNP1236 MTERF4 | SNP1237 PASK | 1 |
| Autism | SNP1081 BCR | SNP1238 PASK | 0.01149158 |
| Autism | SNP1213 SLC39A10 | SNP1238 PASK | 0.02186093 |
| Autism | SNP1225 COL6A3 | SNP1238 PASK | 0.064862587 |
| Autism | SNP1236 MTERF4 | SNP1238 PASK | 0.964783164 |
| Autism | SNP1237 PASK | SNP1238 PASK | 0.964783164 |
| Autism | SNP1081 BCR | SNP1243 ANO7 | 0.012672723 |
| Autism | SNP1155 CD8B | SNP1243 ANO7 | 0.026228981 |
| Autism | SNP1213 SLC39A10 | SNP1243 ANO7 | 0.032516408 |
| Autism | SNP1225 COL6A3 | SNP1243 ANO7 | 0.067753012 |
| Autism | SNP1236 MTERF4 | SNP1243 ANO7 | 0.370762886 |
| Autism | SNP1237 PASK | SNP1243 ANO7 | 0.370762886 |
| Autism | SNP1238 PASK | SNP1243 ANO7 | 0.385072306 |
| Autism | SNP1081 BCR | SNP1246 SEPT2 | 0.010270318 |
| Autism | SNP1155 CD8B | SNP1246 SEPT2 | 0.024727483 |
| Autism | SNP1213 SLC39A10 | SNP1246 SEPT2 | 0.029086154 |
| Autism | SNP1225 COL6A3 | SNP1246 SEPT2 | 0.079094625 |
| Autism | SNP1236 MTERF4 | SNP1246 SEPT2 | 0.356858199 |
| Autism | SNP1237 PASK | SNP1246 SEPT2 | 0.356858199 |
| Autism | SNP1238 PASK | SNP1246 SEPT2 | 0.370645013 |
| Autism | SNP1243 ANO7 | SNP1246 SEPT2 | 0.913284609 |
| Autism | SNP1081 BCR | SNP1248 FARP2 | 0.010270318 |
| Autism | SNP1155 CD8B | SNP1248 FARP2 | 0.024727483 |
| Autism | SNP1213 SLC39A10 | SNP1248 FARP2 | 0.029086154 |
| Autism | SNP1225 COL6A3 | SNP1248 FARP2 | 0.079094625 |
| Autism | SNP1236 MTERF4 | SNP1248 FARP2 | 0.356858199 |
| Autism | SNP1237 PASK | SNP1248 FARP2 | 0.356858199 |
| Autism | SNP1238 PASK | SNP1248 FARP2 | 0.370645013 |
| Autism | SNP1243 ANO7 | SNP1248 FARP2 | 0.913284609 |
| Autism | SNP1246 SEPT2 | SNP1248 FARP2 | 1 |
| Autism | SNP1081 BCR | SNP1249 FARP2 | 0.010021435 |
| Autism | SNP1155 CD8B | SNP1249 FARP2 | 0.024130586 |
| Autism | SNP1213 SLC39A10 | SNP1249 FARP2 | 0.028498977 |
| Autism | SNP1225 COL6A3 | SNP1249 FARP2 | 0.077679117 |
| Autism | SNP1236 MTERF4 | SNP1249 FARP2 | 0.350928487 |
| Autism | SNP1237 PASK | SNP1249 FARP2 | 0.350928487 |
| Autism | SNP1238 PASK | SNP1249 FARP2 | 0.364512878 |
| Autism | SNP1243 ANO7 | SNP1249 FARP2 | 0.898367818 |
| Autism | SNP1246 SEPT2 | SNP1249 FARP2 | 0.983749649 |
| Autism | SNP1248 FARP2 | SNP1249 FARP2 | 0.983749649 |
| Autism | SNP1213 SLC39A10 | SNP1374 PAK2 | 0.038148681 |
| Autism | SNP1225 COL6A3 | SNP1374 PAK2 | 0.011396409 |
| Autism | SNP1236 MTERF4 | SNP1374 PAK2 | 0.017532871 |
| Autism | SNP1237 PASK | SNP1374 PAK2 | 0.017532871 |
| Autism | SNP1238 PASK | SNP1374 PAK2 | 0.018414423 |
| Autism | SNP1243 ANO7 | SNP1374 PAK2 | 0.024982498 |
| Autism | SNP1246 SEPT2 | SNP1374 PAK2 | 0.026283429 |
| Autism | SNP1248 FARP2 | SNP1374 PAK2 | 0.026283429 |
| Autism | SNP1249 FARP2 | SNP1374 PAK2 | 0.025680233 |
| Autism | SNP1155 CD8B | SNP1443 FAT1 | 0.010594317 |
| Autism | SNP1155 CD8B | SNP1444 FAT1 | 0.010721308 |
| Autism | SNP1443 FAT1 | SNP1444 FAT1 | 0.993596904 |
| Autism | SNP1155 CD8B | SNP1445 FAT1 | 0.011779758 |
| Autism | SNP1443 FAT1 | SNP1445 FAT1 | 0.739390788 |
| Autism | SNP1444 FAT1 | SNP1445 FAT1 | 0.744271574 |
| Autism | SNP1155 CD8B | SNP1446 FAT1 | 0.011905005 |
| Autism | SNP1443 FAT1 | SNP1446 FAT1 | 0.733318142 |
| Autism | SNP1444 FAT1 | SNP1446 FAT1 | 0.738159539 |
| Autism | SNP1445 FAT1 | SNP1446 FAT1 | 0.99430376 |
| Autism | SNP1155 CD8B | SNP1447 FAT1 | 0.011905005 |
| Autism | SNP1443 FAT1 | SNP1447 FAT1 | 0.733318142 |
| Autism | SNP1444 FAT1 | SNP1447 FAT1 | 0.738159539 |
| Autism | SNP1445 FAT1 | SNP1447 FAT1 | 0.99430376 |
| Autism | SNP1446 FAT1 | SNP1447 FAT1 | 0.988579694 |
| Autism | SNP1081 BCR | SNP1630 UNC93A | 0.029824013 |
| Autism | SNP1155 CD8B | SNP1630 UNC93A | 0.011371487 |
| Autism | SNP1213 SLC39A10 | SNP1630 UNC93A | 0.016686793 |
| Autism | SNP1225 COL6A3 | SNP1630 UNC93A | 0.092066507 |
| Autism | SNP1236 MTERF4 | SNP1630 UNC93A | 0.025622921 |
| Autism | SNP1237 PASK | SNP1630 UNC93A | 0.025622921 |
| Autism | SNP1238 PASK | SNP1630 UNC93A | 0.026768674 |
| Autism | SNP1243 ANO7 | SNP1630 UNC93A | 0.041029209 |
| Autism | SNP1246 SEPT2 | SNP1630 UNC93A | 0.037979494 |
| Autism | SNP1248 FARP2 | SNP1630 UNC93A | 0.037979494 |
| Autism | SNP1249 FARP2 | SNP1630 UNC93A | 0.037218141 |
| Autism | SNP1374 PAK2 | SNP1630 UNC93A | 0.033409746 |
| Autism | SNP1445 FAT1 | SNP1630 UNC93A | 0.01110653 |
| Autism | SNP1446 FAT1 | SNP1630 UNC93A | 0.011206535 |
| Autism | SNP1447 FAT1 | SNP1630 UNC93A | 0.011206535 |
| Autism | SNP1243 ANO7 | SNP244 LRRK2 | 0.013100913 |
| Autism | SNP1246 SEPT2 | SNP244 LRRK2 | 0.013953927 |
| Autism | SNP1248 FARP2 | SNP244 LRRK2 | 0.013953927 |
| Autism | SNP1249 FARP2 | SNP244 LRRK2 | 0.013522553 |
| Autism | SNP1081 BCR | SNP246 MUC19 | 0.012830233 |
| Autism | SNP1155 CD8B | SNP246 MUC19 | 0.010213 |
| Autism | SNP1213 SLC39A10 | SNP246 MUC19 | 0.017278757 |
| Autism | SNP1236 MTERF4 | SNP246 MUC19 | 0.015147399 |
| Autism | SNP1237 PASK | SNP246 MUC19 | 0.015147399 |
| Autism | SNP1238 PASK | SNP246 MUC19 | 0.015910601 |
| Autism | SNP1243 ANO7 | SNP246 MUC19 | 0.034983392 |
| Autism | SNP1246 SEPT2 | SNP246 MUC19 | 0.036708983 |
| Autism | SNP1248 FARP2 | SNP246 MUC19 | 0.036708983 |
| Autism | SNP1249 FARP2 | SNP246 MUC19 | 0.035928992 |
| Autism | SNP1374 PAK2 | SNP246 MUC19 | 0.01741755 |
| Autism | SNP1630 UNC93A | SNP246 MUC19 | 0.016529917 |
| Autism | SNP244 LRRK2 | SNP246 MUC19 | 0.243179498 |
| Autism | SNP288 UTP20 | SNP289 UTP20 | 1 |
| Autism | SNP288 UTP20 | SNP290 UTP20 | 1 |
| Autism | SNP289 UTP20 | SNP290 UTP20 | 1 |
| Autism | SNP288 UTP20 | SNP291 UTP20 | 0.957861657 |
| Autism | SNP289 UTP20 | SNP291 UTP20 | 0.957861657 |
| Autism | SNP290 UTP20 | SNP291 UTP20 | 0.957861657 |
| Autism | SNP288 UTP20 | SNP292 UTP20 | 0.985886654 |
| Autism | SNP289 UTP20 | SNP292 UTP20 | 0.985886654 |
| Autism | SNP290 UTP20 | SNP292 UTP20 | 0.985886654 |
| Autism | SNP291 UTP20 | SNP292 UTP20 | 0.943581556 |
| Autism | SNP1243 ANO7 | SNP30 PTCHD3 | 0.018184286 |
| Autism | SNP1246 SEPT2 | SNP30 PTCHD3 | 0.019117916 |
| Autism | SNP1248 FARP2 | SNP30 PTCHD3 | 0.019117916 |
| Autism | SNP1249 FARP2 | SNP30 PTCHD3 | 0.018687806 |
| Autism | SNP1374 PAK2 | SNP30 PTCHD3 | 0.012750708 |
| Autism | SNP1445 FAT1 | SNP30 PTCHD3 | 0.023178461 |
| Autism | SNP1446 FAT1 | SNP30 PTCHD3 | 0.023360149 |
| Autism | SNP1447 FAT1 | SNP30 PTCHD3 | 0.023360149 |
| Autism | SNP1630 UNC93A | SNP30 PTCHD3 | 0.013189355 |
| Autism | SNP1155 CD8B | SNP326 SCARB1 | 0.0165783 |
| Autism | SNP1213 SLC39A10 | SNP326 SCARB1 | 0.023392495 |
| Autism | SNP1225 COL6A3 | SNP326 SCARB1 | 0.028813181 |
| Autism | SNP1236 MTERF4 | SNP326 SCARB1 | 0.036804749 |
| Autism | SNP1237 PASK | SNP326 SCARB1 | 0.036804749 |
| Autism | SNP1238 PASK | SNP326 SCARB1 | 0.038426713 |
| Autism | SNP1243 ANO7 | SNP326 SCARB1 | 0.112256764 |
| Autism | SNP1246 SEPT2 | SNP326 SCARB1 | 0.109611401 |
| Autism | SNP1248 FARP2 | SNP326 SCARB1 | 0.109611401 |
| Autism | SNP1249 FARP2 | SNP326 SCARB1 | 0.115107393 |
| Autism | SNP1374 PAK2 | SNP326 SCARB1 | 0.022082457 |
| Autism | SNP244 LRRK2 | SNP326 SCARB1 | 0.025889428 |
| Autism | SNP246 MUC19 | SNP326 SCARB1 | 0.036113083 |
| Autism | SNP1155 CD8B | SNP327 SCARB1 | 0.017357109 |
| Autism | SNP1213 SLC39A10 | SNP327 SCARB1 | 0.011440597 |
| Autism | SNP1225 COL6A3 | SNP327 SCARB1 | 0.02013741 |
| Autism | SNP1236 MTERF4 | SNP327 SCARB1 | 0.01092272 |
| Autism | SNP1237 PASK | SNP327 SCARB1 | 0.01092272 |
| Autism | SNP1238 PASK | SNP327 SCARB1 | 0.011470809 |
| Autism | SNP1243 ANO7 | SNP327 SCARB1 | 0.050298102 |
| Autism | SNP1246 SEPT2 | SNP327 SCARB1 | 0.041974114 |
| Autism | SNP1248 FARP2 | SNP327 SCARB1 | 0.041974114 |
| Autism | SNP1249 FARP2 | SNP327 SCARB1 | 0.041141069 |
| Autism | SNP1374 PAK2 | SNP327 SCARB1 | 0.01149207 |
| Autism | SNP246 MUC19 | SNP327 SCARB1 | 0.012157242 |
| Autism | SNP326 SCARB1 | SNP327 SCARB1 | 0.468123226 |
| Autism | SNP1213 SLC39A10 | SNP33 ARMC4 | 0.027241598 |
| Autism | SNP1243 ANO7 | SNP33 ARMC4 | 0.015464336 |
| Autism | SNP1246 SEPT2 | SNP33 ARMC4 | 0.020371492 |
| Autism | SNP1248 FARP2 | SNP33 ARMC4 | 0.020371492 |
| Autism | SNP1249 FARP2 | SNP33 ARMC4 | 0.019937554 |
| Autism | SNP1374 PAK2 | SNP33 ARMC4 | 0.011150703 |
| Autism | SNP1445 FAT1 | SNP33 ARMC4 | 0.010941149 |
| Autism | SNP1446 FAT1 | SNP33 ARMC4 | 0.011035729 |
| Autism | SNP1447 FAT1 | SNP33 ARMC4 | 0.011035729 |
| Autism | SNP1630 UNC93A | SNP33 ARMC4 | 0.04462629 |
| Autism | SNP246 MUC19 | SNP33 ARMC4 | 0.015531605 |
| Autism | SNP30 PTCHD3 | SNP33 ARMC4 | 0.209018816 |
| Autism | SNP326 SCARB1 | SNP33 ARMC4 | 0.021868656 |
| Autism | SNP326 SCARB1 | SNP377 SCEL | 0.013654337 |
| Autism | SNP327 SCARB1 | SNP377 SCEL | 0.014163443 |
| Autism | SNP1155 CD8B | SNP558 CRAMP1 | 0.022400466 |
| Autism | SNP1213 SLC39A10 | SNP558 CRAMP1 | 0.055238142 |
| Autism | SNP1225 COL6A3 | SNP558 CRAMP1 | 0.029542714 |
| Autism | SNP1236 MTERF4 | SNP558 CRAMP1 | 0.012530072 |
| Autism | SNP1237 PASK | SNP558 CRAMP1 | 0.012530072 |
| Autism | SNP1238 PASK | SNP558 CRAMP1 | 0.013201166 |
| Autism | SNP1243 ANO7 | SNP558 CRAMP1 | 0.038692259 |
| Autism | SNP1246 SEPT2 | SNP558 CRAMP1 | 0.044547551 |
| Autism | SNP1248 FARP2 | SNP558 CRAMP1 | 0.044547551 |
| Autism | SNP1249 FARP2 | SNP558 CRAMP1 | 0.043603009 |
| Autism | SNP1374 PAK2 | SNP558 CRAMP1 | 0.010882195 |
| Autism | SNP1443 FAT1 | SNP558 CRAMP1 | 0.013763925 |
| Autism | SNP1444 FAT1 | SNP558 CRAMP1 | 0.013911822 |
| Autism | SNP1445 FAT1 | SNP558 CRAMP1 | 0.015209805 |
| Autism | SNP1446 FAT1 | SNP558 CRAMP1 | 0.01535487 |
| Autism | SNP1447 FAT1 | SNP558 CRAMP1 | 0.01535487 |
| Autism | SNP1630 UNC93A | SNP558 CRAMP1 | 0.053620918 |
| Autism | SNP244 LRRK2 | SNP558 CRAMP1 | 0.014877857 |
| Autism | SNP246 MUC19 | SNP558 CRAMP1 | 0.015684599 |
| Autism | SNP30 PTCHD3 | SNP558 CRAMP1 | 0.022996919 |
| Autism | SNP326 SCARB1 | SNP558 CRAMP1 | 0.013283198 |
| Autism | SNP327 SCARB1 | SNP558 CRAMP1 | 0.020772976 |
| Autism | SNP33 ARMC4 | SNP558 CRAMP1 | 0.010059194 |
| Autism | SNP1016 SNX21 | SNP584 SLC16A11 | 0.0101933 |
| Autism | SNP1021 MMP9 | SNP584 SLC16A11 | 0.013859162 |
| Autism | SNP1022 SLC12A5 | SNP584 SLC16A11 | 0.013369044 |
| Autism | SNP1023 SLC12A5 | SNP584 SLC16A11 | 0.01128741 |
| Autism | SNP1213 SLC39A10 | SNP584 SLC16A11 | 0.019394498 |
| Autism | SNP1225 COL6A3 | SNP584 SLC16A11 | 0.029564933 |
| Autism | SNP1236 MTERF4 | SNP584 SLC16A11 | 0.02270512 |
| Autism | SNP1237 PASK | SNP584 SLC16A11 | 0.02270512 |
| Autism | SNP1238 PASK | SNP584 SLC16A11 | 0.023844437 |
| Autism | SNP1243 ANO7 | SNP584 SLC16A11 | 0.037362553 |
| Autism | SNP1246 SEPT2 | SNP584 SLC16A11 | 0.039271089 |
| Autism | SNP1248 FARP2 | SNP584 SLC16A11 | 0.039271089 |
| Autism | SNP1249 FARP2 | SNP584 SLC16A11 | 0.041908607 |
| Autism | SNP1374 PAK2 | SNP584 SLC16A11 | 0.015074097 |
| Autism | SNP1630 UNC93A | SNP584 SLC16A11 | 0.03884682 |
| Autism | SNP244 LRRK2 | SNP584 SLC16A11 | 0.016795775 |
| Autism | SNP246 MUC19 | SNP584 SLC16A11 | 0.021585616 |
| Autism | SNP326 SCARB1 | SNP584 SLC16A11 | 0.026400904 |
| Autism | SNP558 CRAMP1 | SNP584 SLC16A11 | 0.035119002 |
| Autism | SNP1021 MMP9 | SNP585 SLC16A11 | 0.012233306 |
| Autism | SNP1022 SLC12A5 | SNP585 SLC16A11 | 0.011788264 |
| Autism | SNP1213 SLC39A10 | SNP585 SLC16A11 | 0.019634499 |
| Autism | SNP1225 COL6A3 | SNP585 SLC16A11 | 0.029906078 |
| Autism | SNP1236 MTERF4 | SNP585 SLC16A11 | 0.022994839 |
| Autism | SNP1237 PASK | SNP585 SLC16A11 | 0.022994839 |
| Autism | SNP1238 PASK | SNP585 SLC16A11 | 0.024144445 |
| Autism | SNP1243 ANO7 | SNP585 SLC16A11 | 0.037847828 |
| Autism | SNP1246 SEPT2 | SNP585 SLC16A11 | 0.039775598 |
| Autism | SNP1248 FARP2 | SNP585 SLC16A11 | 0.039775598 |
| Autism | SNP1249 FARP2 | SNP585 SLC16A11 | 0.042443365 |
| Autism | SNP1374 PAK2 | SNP585 SLC16A11 | 0.015295258 |
| Autism | SNP1630 UNC93A | SNP585 SLC16A11 | 0.039297229 |
| Autism | SNP244 LRRK2 | SNP585 SLC16A11 | 0.017078677 |
| Autism | SNP246 MUC19 | SNP585 SLC16A11 | 0.021874295 |
| Autism | SNP326 SCARB1 | SNP585 SLC16A11 | 0.026734247 |
| Autism | SNP558 CRAMP1 | SNP585 SLC16A11 | 0.035565408 |
| Autism | SNP584 SLC16A11 | SNP585 SLC16A11 | 0.99054192 |
| Autism | SNP1021 MMP9 | SNP586 SLC16A11 | 0.013616972 |
| Autism | SNP1022 SLC12A5 | SNP586 SLC16A11 | 0.013131964 |
| Autism | SNP1023 SLC12A5 | SNP586 SLC16A11 | 0.011077342 |
| Autism | SNP1213 SLC39A10 | SNP586 SLC16A11 | 0.019158893 |
| Autism | SNP1225 COL6A3 | SNP586 SLC16A11 | 0.02922999 |
| Autism | SNP1236 MTERF4 | SNP586 SLC16A11 | 0.02242073 |
| Autism | SNP1237 PASK | SNP586 SLC16A11 | 0.02242073 |
| Autism | SNP1238 PASK | SNP586 SLC16A11 | 0.023549936 |
| Autism | SNP1243 ANO7 | SNP586 SLC16A11 | 0.03688623 |
| Autism | SNP1246 SEPT2 | SNP586 SLC16A11 | 0.038775871 |
| Autism | SNP1248 FARP2 | SNP586 SLC16A11 | 0.038775871 |
| Autism | SNP1249 FARP2 | SNP586 SLC16A11 | 0.041383687 |
| Autism | SNP1374 PAK2 | SNP586 SLC16A11 | 0.014857109 |
| Autism | SNP1630 UNC93A | SNP586 SLC16A11 | 0.038404603 |
| Autism | SNP244 LRRK2 | SNP586 SLC16A11 | 0.016518395 |
| Autism | SNP246 MUC19 | SNP586 SLC16A11 | 0.021302288 |
| Autism | SNP326 SCARB1 | SNP586 SLC16A11 | 0.026073684 |
| Autism | SNP558 CRAMP1 | SNP586 SLC16A11 | 0.034680805 |
| Autism | SNP584 SLC16A11 | SNP586 SLC16A11 | 0.99062295 |
| Autism | SNP585 SLC16A11 | SNP586 SLC16A11 | 0.981254325 |
| Autism | SNP1238 PASK | SNP655 CDH20 | 0.010383727 |
| Autism | SNP1243 ANO7 | SNP655 CDH20 | 0.010809542 |
| Autism | SNP1630 UNC93A | SNP655 CDH20 | 0.010896199 |
| Autism | SNP30 PTCHD3 | SNP655 CDH20 | 0.011768535 |
| Autism | SNP327 SCARB1 | SNP655 CDH20 | 0.010592605 |
| Autism | SNP558 CRAMP1 | SNP655 CDH20 | 0.014821598 |
| Autism | SNP584 SLC16A11 | SNP655 CDH20 | 0.016253324 |
| Autism | SNP585 SLC16A11 | SNP655 CDH20 | 0.016497411 |
| Autism | SNP586 SLC16A11 | SNP655 CDH20 | 0.016013866 |
| Autism | SNP1021 MMP9 | SNP697 XAB2 | 0.010004438 |
| Autism | SNP1081 BCR | SNP697 XAB2 | 0.039105128 |
| Autism | SNP1213 SLC39A10 | SNP697 XAB2 | 0.031106906 |
| Autism | SNP1225 COL6A3 | SNP697 XAB2 | 0.023321139 |
| Autism | SNP1236 MTERF4 | SNP697 XAB2 | 0.016832132 |
| Autism | SNP1237 PASK | SNP697 XAB2 | 0.016832132 |
| Autism | SNP1238 PASK | SNP697 XAB2 | 0.017628468 |
| Autism | SNP1243 ANO7 | SNP697 XAB2 | 0.047945471 |
| Autism | SNP1246 SEPT2 | SNP697 XAB2 | 0.040422228 |
| Autism | SNP1248 FARP2 | SNP697 XAB2 | 0.040422228 |
| Autism | SNP1249 FARP2 | SNP697 XAB2 | 0.039613894 |
| Autism | SNP1630 UNC93A | SNP697 XAB2 | 0.028700222 |
| Autism | SNP246 MUC19 | SNP697 XAB2 | 0.021998241 |
| Autism | SNP326 SCARB1 | SNP697 XAB2 | 0.027794088 |
| Autism | SNP327 SCARB1 | SNP697 XAB2 | 0.022648961 |
| Autism | SNP33 ARMC4 | SNP697 XAB2 | 0.011977172 |
| Autism | SNP558 CRAMP1 | SNP697 XAB2 | 0.03455049 |
| Autism | SNP584 SLC16A11 | SNP697 XAB2 | 0.032449818 |
| Autism | SNP585 SLC16A11 | SNP697 XAB2 | 0.032837078 |
| Autism | SNP586 SLC16A11 | SNP697 XAB2 | 0.032069621 |
| Autism | SNP1021 MMP9 | SNP698 PET100 | 0.010299101 |
| Autism | SNP1081 BCR | SNP698 PET100 | 0.039845151 |
| Autism | SNP1213 SLC39A10 | SNP698 PET100 | 0.03170423 |
| Autism | SNP1225 COL6A3 | SNP698 PET100 | 0.017648748 |
| Autism | SNP1236 MTERF4 | SNP698 PET100 | 0.017195086 |
| Autism | SNP1237 PASK | SNP698 PET100 | 0.017195086 |
| Autism | SNP1238 PASK | SNP698 PET100 | 0.018003964 |
| Autism | SNP1243 ANO7 | SNP698 PET100 | 0.048921393 |
| Autism | SNP1246 SEPT2 | SNP698 PET100 | 0.041260852 |
| Autism | SNP1248 FARP2 | SNP698 PET100 | 0.041260852 |
| Autism | SNP1249 FARP2 | SNP698 PET100 | 0.040440294 |
| Autism | SNP1630 UNC93A | SNP698 PET100 | 0.029268498 |
| Autism | SNP246 MUC19 | SNP698 PET100 | 0.022470221 |
| Autism | SNP326 SCARB1 | SNP698 PET100 | 0.028361253 |
| Autism | SNP327 SCARB1 | SNP698 PET100 | 0.018186383 |
| Autism | SNP33 ARMC4 | SNP698 PET100 | 0.012235438 |
| Autism | SNP558 CRAMP1 | SNP698 PET100 | 0.03526532 |
| Autism | SNP584 SLC16A11 | SNP698 PET100 | 0.033145014 |
| Autism | SNP585 SLC16A11 | SNP698 PET100 | 0.033538293 |
| Autism | SNP586 SLC16A11 | SNP698 PET100 | 0.032758905 |
| Autism | SNP697 XAB2 | SNP698 PET100 | 0.984068459 |
| Autism | SNP1243 ANO7 | SNP763 ZSCAN5B | 0.012212864 |
| Autism | SNP1246 SEPT2 | SNP763 ZSCAN5B | 0.013055489 |
| Autism | SNP1248 FARP2 | SNP763 ZSCAN5B | 0.013055489 |
| Autism | SNP1249 FARP2 | SNP763 ZSCAN5B | 0.012621363 |
| Autism | SNP697 XAB2 | SNP763 ZSCAN5B | 0.017167112 |
| Autism | SNP698 PET100 | SNP763 ZSCAN5B | 0.015175101 |
| Autism | SNP1021 MMP9 | SNP779 C1orf174 | 0.010038646 |
| Autism | SNP1081 BCR | SNP779 C1orf174 | 0.04279898 |
| Autism | SNP1213 SLC39A10 | SNP779 C1orf174 | 0.032489744 |
| Autism | SNP1225 COL6A3 | SNP779 C1orf174 | 0.029688235 |
| Autism | SNP1236 MTERF4 | SNP779 C1orf174 | 0.016730761 |
| Autism | SNP1237 PASK | SNP779 C1orf174 | 0.016730761 |
| Autism | SNP1238 PASK | SNP779 C1orf174 | 0.017499426 |
| Autism | SNP1243 ANO7 | SNP779 C1orf174 | 0.04074972 |
| Autism | SNP1246 SEPT2 | SNP779 C1orf174 | 0.042631641 |
| Autism | SNP1248 FARP2 | SNP779 C1orf174 | 0.042631641 |
| Autism | SNP1249 FARP2 | SNP779 C1orf174 | 0.041809231 |
| Autism | SNP1374 PAK2 | SNP779 C1orf174 | 0.017877962 |
| Autism | SNP1445 FAT1 | SNP779 C1orf174 | 0.011974021 |
| Autism | SNP1446 FAT1 | SNP779 C1orf174 | 0.012075587 |
| Autism | SNP1447 FAT1 | SNP779 C1orf174 | 0.012075587 |
| Autism | SNP1630 UNC93A | SNP779 C1orf174 | 0.024369685 |
| Autism | SNP326 SCARB1 | SNP779 C1orf174 | 0.047816901 |
| Autism | SNP327 SCARB1 | SNP779 C1orf174 | 0.017658402 |
| Autism | SNP558 CRAMP1 | SNP779 C1orf174 | 0.031501924 |
| Autism | SNP584 SLC16A11 | SNP779 C1orf174 | 0.036706621 |
| Autism | SNP585 SLC16A11 | SNP779 C1orf174 | 0.037125253 |
| Autism | SNP586 SLC16A11 | SNP779 C1orf174 | 0.036295591 |
| Autism | SNP697 XAB2 | SNP779 C1orf174 | 0.059399965 |
| Autism | SNP698 PET100 | SNP779 C1orf174 | 0.060480416 |
| Autism | SNP763 ZSCAN5B | SNP779 C1orf174 | 0.180505065 |
| Autism | SNP1021 MMP9 | SNP780 C1orf174 | 0.010038646 |
| Autism | SNP1081 BCR | SNP780 C1orf174 | 0.04279898 |
| Autism | SNP1213 SLC39A10 | SNP780 C1orf174 | 0.032489744 |
| Autism | SNP1225 COL6A3 | SNP780 C1orf174 | 0.029688235 |
| Autism | SNP1236 MTERF4 | SNP780 C1orf174 | 0.016730761 |
| Autism | SNP1237 PASK | SNP780 C1orf174 | 0.016730761 |
| Autism | SNP1238 PASK | SNP780 C1orf174 | 0.017499426 |
| Autism | SNP1243 ANO7 | SNP780 C1orf174 | 0.04074972 |
| Autism | SNP1246 SEPT2 | SNP780 C1orf174 | 0.042631641 |
| Autism | SNP1248 FARP2 | SNP780 C1orf174 | 0.042631641 |
| Autism | SNP1249 FARP2 | SNP780 C1orf174 | 0.041809231 |
| Autism | SNP1374 PAK2 | SNP780 C1orf174 | 0.017877962 |
| Autism | SNP1445 FAT1 | SNP780 C1orf174 | 0.011974021 |
| Autism | SNP1446 FAT1 | SNP780 C1orf174 | 0.012075587 |
| Autism | SNP1447 FAT1 | SNP780 C1orf174 | 0.012075587 |
| Autism | SNP1630 UNC93A | SNP780 C1orf174 | 0.024369685 |
| Autism | SNP326 SCARB1 | SNP780 C1orf174 | 0.047816901 |
| Autism | SNP327 SCARB1 | SNP780 C1orf174 | 0.017658402 |
| Autism | SNP558 CRAMP1 | SNP780 C1orf174 | 0.031501924 |
| Autism | SNP584 SLC16A11 | SNP780 C1orf174 | 0.036706621 |
| Autism | SNP585 SLC16A11 | SNP780 C1orf174 | 0.037125253 |
| Autism | SNP586 SLC16A11 | SNP780 C1orf174 | 0.036295591 |
| Autism | SNP697 XAB2 | SNP780 C1orf174 | 0.059399965 |
| Autism | SNP698 PET100 | SNP780 C1orf174 | 0.060480416 |
| Autism | SNP763 ZSCAN5B | SNP780 C1orf174 | 0.180505065 |
| Autism | SNP779 C1orf174 | SNP780 C1orf174 | 1 |
| Autism | SNP1225 COL6A3 | SNP786 VPS13D | 0.031193821 |
| Autism | SNP1243 ANO7 | SNP786 VPS13D | 0.013320326 |
| Autism | SNP1246 SEPT2 | SNP786 VPS13D | 0.016518532 |
| Autism | SNP1248 FARP2 | SNP786 VPS13D | 0.016518532 |
| Autism | SNP1249 FARP2 | SNP786 VPS13D | 0.016092947 |
| Autism | SNP1374 PAK2 | SNP786 VPS13D | 0.014091135 |
| Autism | SNP1630 UNC93A | SNP786 VPS13D | 0.01451473 |
| Autism | SNP246 MUC19 | SNP786 VPS13D | 0.010358428 |
| Autism | SNP326 SCARB1 | SNP786 VPS13D | 0.010986797 |
| Autism | SNP327 SCARB1 | SNP786 VPS13D | 0.014134388 |
| Autism | SNP33 ARMC4 | SNP786 VPS13D | 0.014655424 |
| Autism | SNP558 CRAMP1 | SNP786 VPS13D | 0.02136078 |
| Autism | SNP584 SLC16A11 | SNP786 VPS13D | 0.029381232 |
| Autism | SNP585 SLC16A11 | SNP786 VPS13D | 0.029770946 |
| Autism | SNP586 SLC16A11 | SNP786 VPS13D | 0.028998731 |
| Autism | SNP779 C1orf174 | SNP786 VPS13D | 0.012902115 |
| Autism | SNP780 C1orf174 | SNP786 VPS13D | 0.012902115 |
| Autism | SNP1155 CD8B | SNP92 BTBD16 | 0.016920525 |
| Autism | SNP1236 MTERF4 | SNP92 BTBD16 | 0.026131006 |
| Autism | SNP1237 PASK | SNP92 BTBD16 | 0.026131006 |
| Autism | SNP1238 PASK | SNP92 BTBD16 | 0.027596835 |
| Autism | SNP1243 ANO7 | SNP92 BTBD16 | 0.033683262 |
| Autism | SNP1246 SEPT2 | SNP92 BTBD16 | 0.033141023 |
| Autism | SNP1248 FARP2 | SNP92 BTBD16 | 0.033141023 |
| Autism | SNP1249 FARP2 | SNP92 BTBD16 | 0.0322469 |
| Autism | SNP1374 PAK2 | SNP92 BTBD16 | 0.01955282 |
| Autism | SNP1630 UNC93A | SNP92 BTBD16 | 0.019537542 |
| Autism | SNP246 MUC19 | SNP92 BTBD16 | 0.015377049 |
| Autism | SNP30 PTCHD3 | SNP92 BTBD16 | 0.01545652 |
| Autism | SNP326 SCARB1 | SNP92 BTBD16 | 0.016534278 |
| Autism | SNP327 SCARB1 | SNP92 BTBD16 | 0.011610836 |
| Autism | SNP558 CRAMP1 | SNP92 BTBD16 | 0.021572919 |
| Autism | SNP584 SLC16A11 | SNP92 BTBD16 | 0.011017287 |
| Autism | SNP585 SLC16A11 | SNP92 BTBD16 | 0.011242134 |
| Autism | SNP586 SLC16A11 | SNP92 BTBD16 | 0.010797112 |
| Autism | SNP697 XAB2 | SNP92 BTBD16 | 0.011451396 |
| Autism | SNP698 PET100 | SNP92 BTBD16 | 0.011799762 |
| Autism | SNP779 C1orf174 | SNP92 BTBD16 | 0.013359919 |
| Autism | SNP780 C1orf174 | SNP92 BTBD16 | 0.013359919 |
| Autism | SNP1213 SLC39A10 | SNP987 VPS16 | 0.014185135 |
| Autism | SNP1236 MTERF4 | SNP987 VPS16 | 0.012838565 |
| Autism | SNP1237 PASK | SNP987 VPS16 | 0.012838565 |
| Autism | SNP1238 PASK | SNP987 VPS16 | 0.013499058 |
| Autism | SNP1243 ANO7 | SNP987 VPS16 | 0.026703546 |
| Autism | SNP1246 SEPT2 | SNP987 VPS16 | 0.024245205 |
| Autism | SNP1248 FARP2 | SNP987 VPS16 | 0.024245205 |
| Autism | SNP1249 FARP2 | SNP987 VPS16 | 0.02369714 |
| Autism | SNP326 SCARB1 | SNP987 VPS16 | 0.010767055 |
| Autism | SNP327 SCARB1 | SNP987 VPS16 | 0.013692409 |
| Autism | SNP558 CRAMP1 | SNP987 VPS16 | 0.025721221 |
| Autism | SNP584 SLC16A11 | SNP987 VPS16 | 0.013975983 |
| Autism | SNP585 SLC16A11 | SNP987 VPS16 | 0.014177321 |
| Autism | SNP586 SLC16A11 | SNP987 VPS16 | 0.013778429 |
| Autism | SNP697 XAB2 | SNP987 VPS16 | 0.015409492 |
| Autism | SNP698 PET100 | SNP987 VPS16 | 0.011884639 |
| Autism | SNP779 C1orf174 | SNP987 VPS16 | 0.020946615 |
| Autism | SNP780 C1orf174 | SNP987 VPS16 | 0.020946615 |
| Autism | SNP786 VPS13D | SNP987 VPS16 | 0.011809427 |
| Autism | SNP92 BTBD16 | SNP987 VPS16 | 0.010883882 |
| Autism | SNP1213 SLC39A10 | SNP988 VPS16 | 0.014185135 |
| Autism | SNP1236 MTERF4 | SNP988 VPS16 | 0.012838565 |
| Autism | SNP1237 PASK | SNP988 VPS16 | 0.012838565 |
| Autism | SNP1238 PASK | SNP988 VPS16 | 0.013499058 |
| Autism | SNP1243 ANO7 | SNP988 VPS16 | 0.026703546 |
| Autism | SNP1246 SEPT2 | SNP988 VPS16 | 0.024245205 |
| Autism | SNP1248 FARP2 | SNP988 VPS16 | 0.024245205 |
| Autism | SNP1249 FARP2 | SNP988 VPS16 | 0.02369714 |
| Autism | SNP326 SCARB1 | SNP988 VPS16 | 0.010767055 |
| Autism | SNP327 SCARB1 | SNP988 VPS16 | 0.013692409 |
| Autism | SNP558 CRAMP1 | SNP988 VPS16 | 0.025721221 |
| Autism | SNP584 SLC16A11 | SNP988 VPS16 | 0.013975983 |
| Autism | SNP585 SLC16A11 | SNP988 VPS16 | 0.014177321 |
| Autism | SNP586 SLC16A11 | SNP988 VPS16 | 0.013778429 |
| Autism | SNP697 XAB2 | SNP988 VPS16 | 0.015409492 |
| Autism | SNP698 PET100 | SNP988 VPS16 | 0.011884639 |
| Autism | SNP779 C1orf174 | SNP988 VPS16 | 0.020946615 |
| Autism | SNP780 C1orf174 | SNP988 VPS16 | 0.020946615 |
| Autism | SNP786 VPS13D | SNP988 VPS16 | 0.011809427 |
| Autism | SNP92 BTBD16 | SNP988 VPS16 | 0.010883882 |
| Autism | SNP987 VPS16 | SNP988 VPS16 | 1 |
| Autism | SNP1213 SLC39A10 | SNP989 PTPRA | 0.015152127 |
| Autism | SNP1236 MTERF4 | SNP989 PTPRA | 0.01377301 |
| Autism | SNP1237 PASK | SNP989 PTPRA | 0.01377301 |
| Autism | SNP1238 PASK | SNP989 PTPRA | 0.01446796 |
| Autism | SNP1243 ANO7 | SNP989 PTPRA | 0.028610806 |
| Autism | SNP1246 SEPT2 | SNP989 PTPRA | 0.025997594 |
| Autism | SNP1248 FARP2 | SNP989 PTPRA | 0.025997594 |
| Autism | SNP1249 FARP2 | SNP989 PTPRA | 0.025421916 |
| Autism | SNP1374 PAK2 | SNP989 PTPRA | 0.010341957 |
| Autism | SNP326 SCARB1 | SNP989 PTPRA | 0.011584408 |
| Autism | SNP327 SCARB1 | SNP989 PTPRA | 0.014661602 |
| Autism | SNP558 CRAMP1 | SNP989 PTPRA | 0.027516741 |
| Autism | SNP584 SLC16A11 | SNP989 PTPRA | 0.015088188 |
| Autism | SNP585 SLC16A11 | SNP989 PTPRA | 0.015300213 |
| Autism | SNP586 SLC16A11 | SNP989 PTPRA | 0.014880127 |
| Autism | SNP697 XAB2 | SNP989 PTPRA | 0.016485182 |
| Autism | SNP698 PET100 | SNP989 PTPRA | 0.012743634 |
| Autism | SNP779 C1orf174 | SNP989 PTPRA | 0.02231133 |
| Autism | SNP780 C1orf174 | SNP989 PTPRA | 0.02231133 |
| Autism | SNP92 BTBD16 | SNP989 PTPRA | 0.01196939 |
| Autism | SNP987 VPS16 | SNP989 PTPRA | 0.916225088 |
| Autism | SNP988 VPS16 | SNP989 PTPRA | 0.916225088 |
| Control | SNP1015 WFDC3 | SNP1016 SNX21 | 0.968277208 |
| Control | SNP1015 WFDC3 | SNP1021 MMP9 | 0.101952057 |
| Control | SNP1016 SNX21 | SNP1021 MMP9 | 0.09819436 |
| Control | SNP1015 WFDC3 | SNP1022 SLC12A5 | 0.065071497 |
| Control | SNP1016 SNX21 | SNP1022 SLC12A5 | 0.062528364 |
| Control | SNP1021 MMP9 | SNP1022 SLC12A5 | 0.832786514 |
| Control | SNP1015 WFDC3 | SNP1023 SLC12A5 | 0.062397886 |
| Control | SNP1016 SNX21 | SNP1023 SLC12A5 | 0.059933517 |
| Control | SNP1021 MMP9 | SNP1023 SLC12A5 | 0.80557797 |
| Control | SNP1022 SLC12A5 | SNP1023 SLC12A5 | 0.903637778 |
| Control | SNP1015 WFDC3 | SNP1081 BCR | 0.029893207 |
| Control | SNP1016 SNX21 | SNP1081 BCR | 0.028944911 |
| Control | SNP1021 MMP9 | SNP1081 BCR | 0.037114846 |
| Control | SNP1022 SLC12A5 | SNP1081 BCR | 0.030908743 |
| Control | SNP1023 SLC12A5 | SNP1081 BCR | 0.029898902 |
| Control | SNP1015 WFDC3 | SNP1213 SLC39A10 | 0.013597898 |
| Control | SNP1016 SNX21 | SNP1213 SLC39A10 | 0.013124161 |
| Control | SNP1021 MMP9 | SNP1225 COL6A3 | 0.010604374 |
| Control | SNP1213 SLC39A10 | SNP1236 MTERF4 | 0.081469235 |
| Control | SNP1213 SLC39A10 | SNP1237 PASK | 0.081469235 |
| Control | SNP1236 MTERF4 | SNP1237 PASK | 1 |
| Control | SNP1213 SLC39A10 | SNP1238 PASK | 0.081469235 |
| Control | SNP1236 MTERF4 | SNP1238 PASK | 1 |
| Control | SNP1237 PASK | SNP1238 PASK | 1 |
| Control | SNP1081 BCR | SNP1243 ANO7 | 0.070128051 |
| Control | SNP1155 CD8B | SNP1243 ANO7 | 0.013251084 |
| Control | SNP1213 SLC39A10 | SNP1243 ANO7 | 0.033724684 |
| Control | SNP1225 COL6A3 | SNP1243 ANO7 | 0.021606421 |
| Control | SNP1236 MTERF4 | SNP1243 ANO7 | 0.184584566 |
| Control | SNP1237 PASK | SNP1243 ANO7 | 0.184584566 |
| Control | SNP1238 PASK | SNP1243 ANO7 | 0.184584566 |
| Control | SNP1081 BCR | SNP1246 SEPT2 | 0.082049486 |
| Control | SNP1155 CD8B | SNP1246 SEPT2 | 0.016244545 |
| Control | SNP1213 SLC39A10 | SNP1246 SEPT2 | 0.039691727 |
| Control | SNP1225 COL6A3 | SNP1246 SEPT2 | 0.025585961 |
| Control | SNP1236 MTERF4 | SNP1246 SEPT2 | 0.341937095 |
| Control | SNP1237 PASK | SNP1246 SEPT2 | 0.341937095 |
| Control | SNP1238 PASK | SNP1246 SEPT2 | 0.341937095 |
| Control | SNP1243 ANO7 | SNP1246 SEPT2 | 0.71558355 |
| Control | SNP1081 BCR | SNP1248 FARP2 | 0.089635854 |
| Control | SNP1155 CD8B | SNP1248 FARP2 | 0.018156518 |
| Control | SNP1213 SLC39A10 | SNP1248 FARP2 | 0.043489218 |
| Control | SNP1225 COL6A3 | SNP1248 FARP2 | 0.028119148 |
| Control | SNP1236 MTERF4 | SNP1248 FARP2 | 0.37376954 |
| Control | SNP1237 PASK | SNP1248 FARP2 | 0.37376954 |
| Control | SNP1238 PASK | SNP1248 FARP2 | 0.37376954 |
| Control | SNP1243 ANO7 | SNP1248 FARP2 | 0.644003464 |
| Control | SNP1246 SEPT2 | SNP1248 FARP2 | 0.915364583 |
| Control | SNP1081 BCR | SNP1249 FARP2 | 0.098739496 |
| Control | SNP1155 CD8B | SNP1249 FARP2 | 0.020456021 |
| Control | SNP1213 SLC39A10 | SNP1249 FARP2 | 0.048046413 |
| Control | SNP1225 COL6A3 | SNP1249 FARP2 | 0.031159521 |
| Control | SNP1236 MTERF4 | SNP1249 FARP2 | 0.261414372 |
| Control | SNP1237 PASK | SNP1249 FARP2 | 0.261414372 |
| Control | SNP1238 PASK | SNP1249 FARP2 | 0.261414372 |
| Control | SNP1243 ANO7 | SNP1249 FARP2 | 0.572738395 |
| Control | SNP1246 SEPT2 | SNP1249 FARP2 | 0.830969267 |
| Control | SNP1248 FARP2 | SNP1249 FARP2 | 0.907801418 |
| Control | SNP1021 MMP9 | SNP1374 PAK2 | 0.021298489 |
| Control | SNP1022 SLC12A5 | SNP1374 PAK2 | 0.017390542 |
| Control | SNP1023 SLC12A5 | SNP1374 PAK2 | 0.016710708 |
| Control | SNP1081 BCR | SNP1374 PAK2 | 0.165499533 |
| Control | SNP1246 SEPT2 | SNP1374 PAK2 | 0.011520939 |
| Control | SNP1248 FARP2 | SNP1374 PAK2 | 0.01278576 |
| Control | SNP1249 FARP2 | SNP1374 PAK2 | 0.014305268 |
| Control | SNP1081 BCR | SNP1443 FAT1 | 0.024988371 |
| Control | SNP1243 ANO7 | SNP1443 FAT1 | 0.022672703 |
| Control | SNP1246 SEPT2 | SNP1443 FAT1 | 0.014059727 |
| Control | SNP1081 BCR | SNP1444 FAT1 | 0.027229908 |
| Control | SNP1243 ANO7 | SNP1444 FAT1 | 0.024921129 |
| Control | SNP1246 SEPT2 | SNP1444 FAT1 | 0.015485067 |
| Control | SNP1443 FAT1 | SNP1444 FAT1 | 0.973435407 |
| Control | SNP1081 BCR | SNP1445 FAT1 | 0.024968294 |
| Control | SNP1236 MTERF4 | SNP1445 FAT1 | 0.013582394 |
| Control | SNP1237 PASK | SNP1445 FAT1 | 0.013582394 |
| Control | SNP1238 PASK | SNP1445 FAT1 | 0.013582394 |
| Control | SNP1243 ANO7 | SNP1445 FAT1 | 0.056978129 |
| Control | SNP1246 SEPT2 | SNP1445 FAT1 | 0.045703811 |
| Control | SNP1248 FARP2 | SNP1445 FAT1 | 0.030678193 |
| Control | SNP1249 FARP2 | SNP1445 FAT1 | 0.017764712 |
| Control | SNP1443 FAT1 | SNP1445 FAT1 | 0.744771123 |
| Control | SNP1444 FAT1 | SNP1445 FAT1 | 0.811654878 |
| Control | SNP1081 BCR | SNP1446 FAT1 | 0.024968294 |
| Control | SNP1236 MTERF4 | SNP1446 FAT1 | 0.013582394 |
| Control | SNP1237 PASK | SNP1446 FAT1 | 0.013582394 |
| Control | SNP1238 PASK | SNP1446 FAT1 | 0.013582394 |
| Control | SNP1243 ANO7 | SNP1446 FAT1 | 0.056978129 |
| Control | SNP1246 SEPT2 | SNP1446 FAT1 | 0.045703811 |
| Control | SNP1248 FARP2 | SNP1446 FAT1 | 0.030678193 |
| Control | SNP1249 FARP2 | SNP1446 FAT1 | 0.017764712 |
| Control | SNP1443 FAT1 | SNP1446 FAT1 | 0.744771123 |
| Control | SNP1444 FAT1 | SNP1446 FAT1 | 0.811654878 |
| Control | SNP1445 FAT1 | SNP1446 FAT1 | 1 |
| Control | SNP1081 BCR | SNP1447 FAT1 | 0.024968294 |
| Control | SNP1236 MTERF4 | SNP1447 FAT1 | 0.013582394 |
| Control | SNP1237 PASK | SNP1447 FAT1 | 0.013582394 |
| Control | SNP1238 PASK | SNP1447 FAT1 | 0.013582394 |
| Control | SNP1243 ANO7 | SNP1447 FAT1 | 0.056978129 |
| Control | SNP1246 SEPT2 | SNP1447 FAT1 | 0.045703811 |
| Control | SNP1248 FARP2 | SNP1447 FAT1 | 0.030678193 |
| Control | SNP1249 FARP2 | SNP1447 FAT1 | 0.017764712 |
| Control | SNP1443 FAT1 | SNP1447 FAT1 | 0.744771123 |
| Control | SNP1444 FAT1 | SNP1447 FAT1 | 0.811654878 |
| Control | SNP1445 FAT1 | SNP1447 FAT1 | 1 |
| Control | SNP1446 FAT1 | SNP1447 FAT1 | 1 |
| Control | SNP1225 COL6A3 | SNP1630 UNC93A | 0.053464662 |
| Control | SNP1246 SEPT2 | SNP1630 UNC93A | 0.011520939 |
| Control | SNP1248 FARP2 | SNP1630 UNC93A | 0.01278576 |
| Control | SNP1249 FARP2 | SNP1630 UNC93A | 0.014305268 |
| Control | SNP1225 COL6A3 | SNP244 LRRK2 | 0.010130364 |
| Control | SNP1243 ANO7 | SNP244 LRRK2 | 0.017126745 |
| Control | SNP1246 SEPT2 | SNP244 LRRK2 | 0.021160753 |
| Control | SNP1249 FARP2 | SNP244 LRRK2 | 0.01027483 |
| Control | SNP1155 CD8B | SNP246 MUC19 | 0.023272207 |
| Control | SNP1225 COL6A3 | SNP246 MUC19 | 0.03487614 |
| Control | SNP1243 ANO7 | SNP246 MUC19 | 0.027268542 |
| Control | SNP1246 SEPT2 | SNP246 MUC19 | 0.032604619 |
| Control | SNP244 LRRK2 | SNP246 MUC19 | 0.138760864 |
| Control | SNP288 UTP20 | SNP289 UTP20 | 1 |
| Control | SNP288 UTP20 | SNP290 UTP20 | 1 |
| Control | SNP289 UTP20 | SNP290 UTP20 | 1 |
| Control | SNP288 UTP20 | SNP291 UTP20 | 1 |
| Control | SNP289 UTP20 | SNP291 UTP20 | 1 |
| Control | SNP290 UTP20 | SNP291 UTP20 | 1 |
| Control | SNP288 UTP20 | SNP292 UTP20 | 1 |
| Control | SNP289 UTP20 | SNP292 UTP20 | 1 |
| Control | SNP290 UTP20 | SNP292 UTP20 | 1 |
| Control | SNP291 UTP20 | SNP292 UTP20 | 1 |
| Control | SNP1236 MTERF4 | SNP30 PTCHD3 | 0.080366303 |
| Control | SNP1237 PASK | SNP30 PTCHD3 | 0.080366303 |
| Control | SNP1238 PASK | SNP30 PTCHD3 | 0.080366303 |
| Control | SNP1243 ANO7 | SNP30 PTCHD3 | 0.076147537 |
| Control | SNP1246 SEPT2 | SNP30 PTCHD3 | 0.089620624 |
| Control | SNP1248 FARP2 | SNP30 PTCHD3 | 0.042267769 |
| Control | SNP1249 FARP2 | SNP30 PTCHD3 | 0.046837957 |
| Control | SNP246 MUC19 | SNP30 PTCHD3 | 0.012097667 |
| Control | SNP1015 WFDC3 | SNP326 SCARB1 | 0.020787794 |
| Control | SNP1016 SNX21 | SNP326 SCARB1 | 0.020028775 |
| Control | SNP1081 BCR | SNP326 SCARB1 | 0.198879552 |
| Control | SNP1243 ANO7 | SNP326 SCARB1 | 0.053088861 |
| Control | SNP1246 SEPT2 | SNP326 SCARB1 | 0.062672383 |
| Control | SNP1248 FARP2 | SNP326 SCARB1 | 0.01584507 |
| Control | SNP1249 FARP2 | SNP326 SCARB1 | 0.017669563 |
| Control | SNP1374 PAK2 | SNP326 SCARB1 | 0.031071005 |
| Control | SNP1443 FAT1 | SNP326 SCARB1 | 0.017376961 |
| Control | SNP1444 FAT1 | SNP326 SCARB1 | 0.019029361 |
| Control | SNP1445 FAT1 | SNP326 SCARB1 | 0.017191177 |
| Control | SNP1446 FAT1 | SNP326 SCARB1 | 0.017191177 |
| Control | SNP1447 FAT1 | SNP326 SCARB1 | 0.017191177 |
| Control | SNP244 LRRK2 | SNP326 SCARB1 | 0.025430739 |
| Control | SNP246 MUC19 | SNP326 SCARB1 | 0.085040143 |
| Control | SNP1081 BCR | SNP327 SCARB1 | 0.165499533 |
| Control | SNP1246 SEPT2 | SNP327 SCARB1 | 0.011520939 |
| Control | SNP1248 FARP2 | SNP327 SCARB1 | 0.01278576 |
| Control | SNP1249 FARP2 | SNP327 SCARB1 | 0.014305268 |
| Control | SNP1374 PAK2 | SNP327 SCARB1 | 0.025476782 |
| Control | SNP246 MUC19 | SNP327 SCARB1 | 0.016164349 |
| Control | SNP326 SCARB1 | SNP327 SCARB1 | 0.530331022 |
| Control | SNP1445 FAT1 | SNP377 SCEL | 0.015483894 |
| Control | SNP1446 FAT1 | SNP377 SCEL | 0.015483894 |
| Control | SNP1447 FAT1 | SNP377 SCEL | 0.015483894 |
| Control | SNP1155 CD8B | SNP558 CRAMP1 | 0.010155982 |
| Control | SNP1243 ANO7 | SNP558 CRAMP1 | 0.011941119 |
| Control | SNP1246 SEPT2 | SNP558 CRAMP1 | 0.01432576 |
| Control | SNP1248 FARP2 | SNP558 CRAMP1 | 0.01584507 |
| Control | SNP1249 FARP2 | SNP558 CRAMP1 | 0.017669563 |
| Control | SNP1630 UNC93A | SNP558 CRAMP1 | 0.031071005 |
| Control | SNP1155 CD8B | SNP584 SLC16A11 | 0.025293791 |
| Control | SNP1225 COL6A3 | SNP584 SLC16A11 | 0.016511921 |
| Control | SNP1236 MTERF4 | SNP584 SLC16A11 | 0.033163575 |
| Control | SNP1237 PASK | SNP584 SLC16A11 | 0.033163575 |
| Control | SNP1238 PASK | SNP584 SLC16A11 | 0.033163575 |
| Control | SNP1243 ANO7 | SNP584 SLC16A11 | 0.056129598 |
| Control | SNP1246 SEPT2 | SNP584 SLC16A11 | 0.066893243 |
| Control | SNP1248 FARP2 | SNP584 SLC16A11 | 0.016098185 |
| Control | SNP1249 FARP2 | SNP584 SLC16A11 | 0.018137002 |
| Control | SNP1445 FAT1 | SNP584 SLC16A11 | 0.027997027 |
| Control | SNP1446 FAT1 | SNP584 SLC16A11 | 0.027997027 |
| Control | SNP1447 FAT1 | SNP584 SLC16A11 | 0.027997027 |
| Control | SNP1630 UNC93A | SNP584 SLC16A11 | 0.078240858 |
| Control | SNP246 MUC19 | SNP584 SLC16A11 | 0.049956746 |
| Control | SNP326 SCARB1 | SNP584 SLC16A11 | 0.040699245 |
| Control | SNP1155 CD8B | SNP585 SLC16A11 | 0.025293791 |
| Control | SNP1225 COL6A3 | SNP585 SLC16A11 | 0.016511921 |
| Control | SNP1236 MTERF4 | SNP585 SLC16A11 | 0.033163575 |
| Control | SNP1237 PASK | SNP585 SLC16A11 | 0.033163575 |
| Control | SNP1238 PASK | SNP585 SLC16A11 | 0.033163575 |
| Control | SNP1243 ANO7 | SNP585 SLC16A11 | 0.056129598 |
| Control | SNP1246 SEPT2 | SNP585 SLC16A11 | 0.066893243 |
| Control | SNP1248 FARP2 | SNP585 SLC16A11 | 0.016098185 |
| Control | SNP1249 FARP2 | SNP585 SLC16A11 | 0.018137002 |
| Control | SNP1445 FAT1 | SNP585 SLC16A11 | 0.027997027 |
| Control | SNP1446 FAT1 | SNP585 SLC16A11 | 0.027997027 |
| Control | SNP1447 FAT1 | SNP585 SLC16A11 | 0.027997027 |
| Control | SNP1630 UNC93A | SNP585 SLC16A11 | 0.078240858 |
| Control | SNP246 MUC19 | SNP585 SLC16A11 | 0.049956746 |
| Control | SNP326 SCARB1 | SNP585 SLC16A11 | 0.040699245 |
| Control | SNP584 SLC16A11 | SNP585 SLC16A11 | 1 |
| Control | SNP1155 CD8B | SNP586 SLC16A11 | 0.025293791 |
| Control | SNP1225 COL6A3 | SNP586 SLC16A11 | 0.016511921 |
| Control | SNP1236 MTERF4 | SNP586 SLC16A11 | 0.033163575 |
| Control | SNP1237 PASK | SNP586 SLC16A11 | 0.033163575 |
| Control | SNP1238 PASK | SNP586 SLC16A11 | 0.033163575 |
| Control | SNP1243 ANO7 | SNP586 SLC16A11 | 0.056129598 |
| Control | SNP1246 SEPT2 | SNP586 SLC16A11 | 0.066893243 |
| Control | SNP1248 FARP2 | SNP586 SLC16A11 | 0.016098185 |
| Control | SNP1249 FARP2 | SNP586 SLC16A11 | 0.018137002 |
| Control | SNP1445 FAT1 | SNP586 SLC16A11 | 0.027997027 |
| Control | SNP1446 FAT1 | SNP586 SLC16A11 | 0.027997027 |
| Control | SNP1447 FAT1 | SNP586 SLC16A11 | 0.027997027 |
| Control | SNP1630 UNC93A | SNP586 SLC16A11 | 0.078240858 |
| Control | SNP246 MUC19 | SNP586 SLC16A11 | 0.049956746 |
| Control | SNP326 SCARB1 | SNP586 SLC16A11 | 0.040699245 |
| Control | SNP584 SLC16A11 | SNP586 SLC16A11 | 1 |
| Control | SNP585 SLC16A11 | SNP586 SLC16A11 | 1 |
| Control | SNP1213 SLC39A10 | SNP655 CDH20 | 0.031536939 |
| Control | SNP1015 WFDC3 | SNP697 XAB2 | 0.013597898 |
| Control | SNP1016 SNX21 | SNP697 XAB2 | 0.013124161 |
| Control | SNP1213 SLC39A10 | SNP697 XAB2 | 0.248599443 |
| Control | SNP1236 MTERF4 | SNP697 XAB2 | 0.081469235 |
| Control | SNP1237 PASK | SNP697 XAB2 | 0.081469235 |
| Control | SNP1238 PASK | SNP697 XAB2 | 0.081469235 |
| Control | SNP1243 ANO7 | SNP697 XAB2 | 0.140452815 |
| Control | SNP1246 SEPT2 | SNP697 XAB2 | 0.164329126 |
| Control | SNP1248 FARP2 | SNP697 XAB2 | 0.043489218 |
| Control | SNP1249 FARP2 | SNP697 XAB2 | 0.048046413 |
| Control | SNP1443 FAT1 | SNP697 XAB2 | 0.011366773 |
| Control | SNP1444 FAT1 | SNP697 XAB2 | 0.012426219 |
| Control | SNP1445 FAT1 | SNP697 XAB2 | 0.011284489 |
| Control | SNP1446 FAT1 | SNP697 XAB2 | 0.011284489 |
| Control | SNP1447 FAT1 | SNP697 XAB2 | 0.011284489 |
| Control | SNP244 LRRK2 | SNP697 XAB2 | 0.016494198 |
| Control | SNP246 MUC19 | SNP697 XAB2 | 0.053616544 |
| Control | SNP326 SCARB1 | SNP697 XAB2 | 0.098181657 |
| Control | SNP584 SLC16A11 | SNP697 XAB2 | 0.108653067 |
| Control | SNP585 SLC16A11 | SNP697 XAB2 | 0.108653067 |
| Control | SNP655 CDH20 | SNP697 XAB2 | 0.031536939 |
| Control | SNP1015 WFDC3 | SNP698 PET100 | 0.013597898 |
| Control | SNP1016 SNX21 | SNP698 PET100 | 0.013124161 |
| Control | SNP1213 SLC39A10 | SNP698 PET100 | 0.248599443 |
| Control | SNP1236 MTERF4 | SNP698 PET100 | 0.081469235 |
| Control | SNP1237 PASK | SNP698 PET100 | 0.081469235 |
| Control | SNP1238 PASK | SNP698 PET100 | 0.081469235 |
| Control | SNP1243 ANO7 | SNP698 PET100 | 0.140452815 |
| Control | SNP1246 SEPT2 | SNP698 PET100 | 0.164329126 |
| Control | SNP1248 FARP2 | SNP698 PET100 | 0.043489218 |
| Control | SNP1249 FARP2 | SNP698 PET100 | 0.048046413 |
| Control | SNP1443 FAT1 | SNP698 PET100 | 0.011366773 |
| Control | SNP1444 FAT1 | SNP698 PET100 | 0.012426219 |
| Control | SNP1445 FAT1 | SNP698 PET100 | 0.011284489 |
| Control | SNP1446 FAT1 | SNP698 PET100 | 0.011284489 |
| Control | SNP1447 FAT1 | SNP698 PET100 | 0.011284489 |
| Control | SNP244 LRRK2 | SNP698 PET100 | 0.016494198 |
| Control | SNP246 MUC19 | SNP698 PET100 | 0.053616544 |
| Control | SNP326 SCARB1 | SNP698 PET100 | 0.098181657 |
| Control | SNP584 SLC16A11 | SNP698 PET100 | 0.108653067 |
| Control | SNP585 SLC16A11 | SNP698 PET100 | 0.108653067 |
| Control | SNP655 CDH20 | SNP698 PET100 | 0.031536939 |
| Control | SNP697 XAB2 | SNP698 PET100 | 1 |
| Control | SNP1630 UNC93A | SNP763 ZSCAN5B | 0.026314974 |
| Control | SNP327 SCARB1 | SNP763 ZSCAN5B | 0.026314974 |
| Control | SNP1015 WFDC3 | SNP779 C1orf174 | 0.059870266 |
| Control | SNP1016 SNX21 | SNP779 C1orf174 | 0.057971014 |
| Control | SNP1243 ANO7 | SNP779 C1orf174 | 0.033724684 |
| Control | SNP1246 SEPT2 | SNP779 C1orf174 | 0.039691727 |
| Control | SNP1443 FAT1 | SNP779 C1orf174 | 0.011366773 |
| Control | SNP1444 FAT1 | SNP779 C1orf174 | 0.012426219 |
| Control | SNP1445 FAT1 | SNP779 C1orf174 | 0.011284489 |
| Control | SNP1446 FAT1 | SNP779 C1orf174 | 0.011284489 |
| Control | SNP1447 FAT1 | SNP779 C1orf174 | 0.011284489 |
| Control | SNP244 LRRK2 | SNP779 C1orf174 | 0.016494198 |
| Control | SNP246 MUC19 | SNP779 C1orf174 | 0.053616544 |
| Control | SNP326 SCARB1 | SNP779 C1orf174 | 0.098181657 |
| Control | SNP584 SLC16A11 | SNP779 C1orf174 | 0.108653067 |
| Control | SNP585 SLC16A11 | SNP779 C1orf174 | 0.108653067 |
| Control | SNP697 XAB2 | SNP779 C1orf174 | 0.248599443 |
| Control | SNP698 PET100 | SNP779 C1orf174 | 0.248599443 |
| Control | SNP1015 WFDC3 | SNP780 C1orf174 | 0.059870266 |
| Control | SNP1016 SNX21 | SNP780 C1orf174 | 0.057971014 |
| Control | SNP1243 ANO7 | SNP780 C1orf174 | 0.033724684 |
| Control | SNP1246 SEPT2 | SNP780 C1orf174 | 0.039691727 |
| Control | SNP1443 FAT1 | SNP780 C1orf174 | 0.011366773 |
| Control | SNP1444 FAT1 | SNP780 C1orf174 | 0.012426219 |
| Control | SNP1445 FAT1 | SNP780 C1orf174 | 0.011284489 |
| Control | SNP1446 FAT1 | SNP780 C1orf174 | 0.011284489 |
| Control | SNP1447 FAT1 | SNP780 C1orf174 | 0.011284489 |
| Control | SNP244 LRRK2 | SNP780 C1orf174 | 0.016494198 |
| Control | SNP246 MUC19 | SNP780 C1orf174 | 0.053616544 |
| Control | SNP326 SCARB1 | SNP780 C1orf174 | 0.098181657 |
| Control | SNP584 SLC16A11 | SNP780 C1orf174 | 0.108653067 |
| Control | SNP585 SLC16A11 | SNP780 C1orf174 | 0.108653067 |
| Control | SNP697 XAB2 | SNP780 C1orf174 | 0.248599443 |
| Control | SNP698 PET100 | SNP780 C1orf174 | 0.248599443 |
| Control | SNP779 C1orf174 | SNP780 C1orf174 | 1 |
| Control | SNP1213 SLC39A10 | SNP786 VPS13D | 0.036478624 |
| Control | SNP1236 MTERF4 | SNP786 VPS13D | 0.010452036 |
| Control | SNP1237 PASK | SNP786 VPS13D | 0.010452036 |
| Control | SNP1238 PASK | SNP786 VPS13D | 0.010452036 |
| Control | SNP697 XAB2 | SNP786 VPS13D | 0.036478624 |
| Control | SNP698 PET100 | SNP786 VPS13D | 0.036478624 |
| Control | SNP1236 MTERF4 | SNP987 VPS16 | 0.025476782 |
| Control | SNP1237 PASK | SNP987 VPS16 | 0.025476782 |
| Control | SNP1238 PASK | SNP987 VPS16 | 0.025476782 |
| Control | SNP1243 ANO7 | SNP987 VPS16 | 0.101744852 |
| Control | SNP1246 SEPT2 | SNP987 VPS16 | 0.119746974 |
| Control | SNP1248 FARP2 | SNP987 VPS16 | 0.056476258 |
| Control | SNP1249 FARP2 | SNP987 VPS16 | 0.062582734 |
| Control | SNP1445 FAT1 | SNP987 VPS16 | 0.013582394 |
| Control | SNP1446 FAT1 | SNP987 VPS16 | 0.013582394 |
| Control | SNP1447 FAT1 | SNP987 VPS16 | 0.013582394 |
| Control | SNP1630 UNC93A | SNP987 VPS16 | 0.025476782 |
| Control | SNP246 MUC19 | SNP987 VPS16 | 0.016164349 |
| Control | SNP30 PTCHD3 | SNP987 VPS16 | 0.183950768 |
| Control | SNP326 SCARB1 | SNP987 VPS16 | 0.031071005 |
| Control | SNP558 CRAMP1 | SNP987 VPS16 | 0.031071005 |
| Control | SNP585 SLC16A11 | SNP987 VPS16 | 0.078240858 |
| Control | SNP697 XAB2 | SNP987 VPS16 | 0.081469235 |
| Control | SNP698 PET100 | SNP987 VPS16 | 0.081469235 |
| Control | SNP779 C1orf174 | SNP987 VPS16 | 0.081469235 |
| Control | SNP780 C1orf174 | SNP987 VPS16 | 0.081469235 |
| Control | SNP1236 MTERF4 | SNP988 VPS16 | 0.025476782 |
| Control | SNP1237 PASK | SNP988 VPS16 | 0.025476782 |
| Control | SNP1238 PASK | SNP988 VPS16 | 0.025476782 |
| Control | SNP1243 ANO7 | SNP988 VPS16 | 0.101744852 |
| Control | SNP1246 SEPT2 | SNP988 VPS16 | 0.119746974 |
| Control | SNP1248 FARP2 | SNP988 VPS16 | 0.056476258 |
| Control | SNP1249 FARP2 | SNP988 VPS16 | 0.062582734 |
| Control | SNP1445 FAT1 | SNP988 VPS16 | 0.013582394 |
| Control | SNP1446 FAT1 | SNP988 VPS16 | 0.013582394 |
| Control | SNP1447 FAT1 | SNP988 VPS16 | 0.013582394 |
| Control | SNP1630 UNC93A | SNP988 VPS16 | 0.025476782 |
| Control | SNP246 MUC19 | SNP988 VPS16 | 0.016164349 |
| Control | SNP30 PTCHD3 | SNP988 VPS16 | 0.183950768 |
| Control | SNP326 SCARB1 | SNP988 VPS16 | 0.031071005 |
| Control | SNP558 CRAMP1 | SNP988 VPS16 | 0.031071005 |
| Control | SNP585 SLC16A11 | SNP988 VPS16 | 0.078240858 |
| Control | SNP697 XAB2 | SNP988 VPS16 | 0.081469235 |
| Control | SNP698 PET100 | SNP988 VPS16 | 0.081469235 |
| Control | SNP779 C1orf174 | SNP988 VPS16 | 0.081469235 |
| Control | SNP780 C1orf174 | SNP988 VPS16 | 0.081469235 |
| Control | SNP987 VPS16 | SNP988 VPS16 | 1 |
| Control | SNP1236 MTERF4 | SNP989 PTPRA | 0.025476782 |
| Control | SNP1237 PASK | SNP989 PTPRA | 0.025476782 |
| Control | SNP1238 PASK | SNP989 PTPRA | 0.025476782 |
| Control | SNP1243 ANO7 | SNP989 PTPRA | 0.101744852 |
| Control | SNP1246 SEPT2 | SNP989 PTPRA | 0.119746974 |
| Control | SNP1248 FARP2 | SNP989 PTPRA | 0.056476258 |
| Control | SNP1249 FARP2 | SNP989 PTPRA | 0.062582734 |
| Control | SNP1445 FAT1 | SNP989 PTPRA | 0.013582394 |
| Control | SNP1446 FAT1 | SNP989 PTPRA | 0.013582394 |
| Control | SNP1447 FAT1 | SNP989 PTPRA | 0.013582394 |
| Control | SNP1630 UNC93A | SNP989 PTPRA | 0.025476782 |
| Control | SNP246 MUC19 | SNP989 PTPRA | 0.016164349 |
| Control | SNP30 PTCHD3 | SNP989 PTPRA | 0.183950768 |
| Control | SNP326 SCARB1 | SNP989 PTPRA | 0.031071005 |
| Control | SNP558 CRAMP1 | SNP989 PTPRA | 0.031071005 |
| Control | SNP585 SLC16A11 | SNP989 PTPRA | 0.078240858 |
| Control | SNP697 XAB2 | SNP989 PTPRA | 0.081469235 |
| Control | SNP698 PET100 | SNP989 PTPRA | 0.081469235 |
| Control | SNP779 C1orf174 | SNP989 PTPRA | 0.081469235 |
| Control | SNP780 C1orf174 | SNP989 PTPRA | 0.081469235 |
| Control | SNP987 VPS16 | SNP989 PTPRA | 1 |
| Control | SNP988 VPS16 | SNP989 PTPRA | 1 |
| Control | SNP33 ARMC4 |  |  |
| Control | SNP92 BTBD16 |  |  |
